# Two mechanisms for accelerated diffusion of COVID-19 outbreaks in regions with high intensity of population and polluting industrialization: the air pollution-to-human and human-to-human transmission dynamics

**DOI:** 10.1101/2020.04.06.20055657

**Authors:** Mario Coccia

## Abstract

**What is COVID-19?:** Coronavirus disease 2019 (COVID-19) is viral infection that generates a severe acute respiratory syndrome with serious pneumonia that may result in progressive respiratory failure and death.

**What are the goals of this investigation?:** This study explains the geo-environmental determinants of the accelerated diffusion of COVID-19 in Italy that is generating a high level of deaths and suggests general lessons learned for a strategy to cope with future epidemics similar to COVID-19 to reduce viral infectivity and negative impacts in economic systems and society.

**What are the results of this study?:** The main results are:

- The accelerate and vast diffusion of COVID-19 in North Italy has a high association with air pollution.
- Hinterland cities have average days of exceeding the limits set for PM_10_ (particulate matter 10 micrometers or less in diameter) equal to 80 days, and an average number of infected more than 2,000 individuals as of April 1^st^, 2020, coastal cities have days of exceeding the limits set for PM_10_ equal to 60 days and have about 700 infected in average.
- Cities that average number of 125 days exceeding the limits set for PM_10_, last year, they have an average number of infected individual higher than 3,200 units, whereas cities having less than 100 days (average number of 48 days) exceeding the limits set for PM_10_, they have an average number of about 900 infected individuals.
- *The results reveal that accelerated transmission dynamics of COVID-19 in specific environments is due to two mechanisms given by: air pollution-to-human transmission and human-to-human transmission; in particular, the mechanisms of air pollution-to-human transmission play a critical role rather than human-to-human transmission*.
- The finding here suggests that to minimize future epidemic similar to COVID-19, the max number of days per year in which cities can exceed the limits set for PM_10_ or for ozone, considering their meteorological condition, is less than 50 days. After this critical threshold, the analytical output here suggests that environmental inconsistencies because of the combination between air pollution and meteorological conditions (with high moisture%, low wind speed and fog) trigger a take-off of viral infectivity (accelerated epidemic diffusion) with damages for health of population, economy and society.

**What is a socioeconomic strategy to prevent future epidemics similar to COVID-19?:** Considering the complex interaction between air pollution, meteorological conditions and biological characteristics of viral infectivity, lessons learned for COVID-19 have to be applied for a proactive socioeconomic strategy to cope with future epidemics, especially an environmental policy based on reduction of air pollution mainly in hinterland zones of countries, having low wind speed, high percentage of moisture and fog that create an environment that can damage immune system of people and foster a fast transmission of viral infectivity similar to the COVID-19.

This study must conclude that a strategy to prevent future epidemics similar to COVID 19 has also to be designed in environmental and sustainability science and not only in terms of biology.

## INTRODUCTION

This study has two goals. The first is to explain the main factors determining the diffusion of COVID-19 that is generating a high level of deaths. The second is to suggest a strategy to cope with future epidemic threats with of accelerated viral infectivity in society.

Coronavirus disease 2019 (COVID-19) is viral infection that generates a severe acute respiratory syndrome with serious clinical symptoms given by fever, dry cough, dyspnea, and pneumonia and may result in progressive respiratory failure and death. Kucharski et al. (2020) argue that COVID-19 transmission declined in Wuhan (China) during late January, 2020 (WHO, 2019, 2020, 2020a; nCoV-2019 Data Working Group, 2020). However, as more infected individuals arrive in international locations before control measures are applied, numerous epidemic chains have led to new outbreaks in different nations worldwide (Xu and Kraemer Moritz, 2020; Wang et al., 2020; Wu et al., 2020). An outbreak of COVID-19 has led to more than 13,900 confirmed deaths in Italy and more than 51,000 deaths worldwide as of April 1^st^, 2020 (Johns Hopkins Center for System Science and Engineering, 2020; cf., Dong et al., 2020). Understanding the prime factors of transmission dynamics of COVID-19 in Italy, having the highest number of deaths worldwide, is crucial for explaining possible relationships underlying the temporal and spatial aspects of the diffusion of this viral infectivity. Results here are basic to design a strategy to prevent future epidemics similar to COVID-19 that generates health and socioeconomic issues for nations and globally.

Currently, as people with the COVID-19 infection arrive in countries or regions with low ongoing transmission, efforts should be done to stop transmission, prevent potential outbreaks and to avoid second and subsequent waves of a COVID-19 epidemic (European Centre for Disease Prevention and Control, 2020; Quilty and Clifford, 2020; Wells et al., 2020). Wells et al. (2020) argue that at the very early stage of the epidemic, reduction in the rate of exportation could delay the importation of cases into cities or nations unaffected by the COVID-19, to gain time to coordinate an appropriate public health response. After that, rapid contact tracing is basic within the epicenter and within and between importation cities to limit human-to-human transmission outside of outbreak countries, also applying appropriate isolation of cases (Wells et al., 2020). The case of severe acute respiratory syndrome outbreak in 2003 started in southern China was able to be controlled through tracing contacts of cases because the majority of transmission occurred after symptom onset (Glasser et al., 2011). These interventions also play a critical role in response to outbreaks where onset of symptoms and infectiousness are concurrent, such as Ebola virus disease (WHO, 2020b; Swanson et al., 2018), MERS (Public Health England, 2019; Kang et al., 2016) and other viral diseases (Hoang et al., 2019; European Centre for Disease Prevention and Control. 2020a). Kucharski, et al. (2020) claim that the isolation of cases and contact tracing can be less effective for COVID-19 because infectiousness starts before the onset of symptoms (cf., Fraser et al., 2004; Peak et al., 2017). Hellewell et al. (2020) show that effective contact tracing and case isolation is enough to control a new outbreak of COVID-19 within 3 months, but the probability of control decreases with long delays from symptom onset to isolation that increase transmission before symptoms. However, it is unclear if these efforts will achieve the control of transmission of COVID-19. In the presence of COVID19 outbreaks, it is crucial to understand the determinants of the transmission dynamics of this viral infectious disease for designing strategies to stop or reduce diffusion, empowering health policy with economic, social and environmental policies. This study focuses on statistical analyses of association between infected people and environmental, demographic and geographical factors that can explain transmission dynamics over time, and provide insights into the environmental situation to prevent and apply, *a priori*, appropriate control measures (Camacho. Et al., 2015; Funk et al., 2017; Riley et al., 2003). In particular, this study here can explain, whenever possible, factors determining the accelerated viral infectivity in specific regions to guide policymakers to prevent future epidemics similar to COVID-19 (Cooper et al., 2006; Kucharski et al., 2015). However, there are several challenges to such studies, particularly in real time. Sources may be biased, incomplete, or only capture certain aspects of the on-going outbreak dynamics.

## DATA AND STUDY DESIGN

The complex problem of viral infectivity of COVID-19 is analyzed here in a perspective of reductionist approach, considering the geo-environmental and demographic factors that we study to explain the relationships supporting the transmission dynamics (cf., Linstone, 1999). In addition, the investigation of the causes of the accelerated diffusion of viral infectivity is done with a philosophical approach *sensu* the philosopher Vico^1^ (Flint, 1884). In particular, the method of inquiry is also based on Kantian approach in which theoretical framework and empirical data complement each other and are inseparable. In this case the truth on this phenomenon, transmission dynamics of COVID-19, is a result of synthesis (Churchman, 1971).

### 1.1 Data and their sources

This study focuses on *N*=55 Italian cities that are provincial capitals. Sources of data are The Ministry of Health in Italy for epidemiological data (Ministero della Salute, 2020), Legambiente (2019) for data of air pollution deriving from the Regional Agencies for Environmental Protection in Italy, il Meteo (2020) for data of weather trend based on meteorological stations of Italian province capitals, The Italian National Institute of Statistics for density of population concerning cities under study (ISTAT, 2020).

### 1.2 Measures

The unit of analysis is main Italian provincial cities. In a perspective of reductionism approach for statistical analysis and decision making, this study focuses on the following measures.

▪ Pollution: total days exceeding the limits set for PM_10_ (particulate matter 10 micrometers or less in diameter) or for ozone in the 55 Italian provincial capitals over 2018. This measure is stable over time and the strategy of using the year 2018, before the COVID-19 outbreak in Italy, is to include the health effects of exposures to pollutants, such as airborne particulate matter and ozone (Brunekreef et al., 2002). In fact, days of air pollution within Italian cities are a main factor that has affected health of population and environment (Legambiente, 2019).
▪ Diffusion of COVID19. Number of infected from 17 March, 2020 to April 2020 (Ministero della Salute, 2020). Infected are detected with COVID-19 tests according to following criteria:
  − Have fever or lower respiratory symptoms (cough, shortness of breath) and close contact with a confirmed COVID-19 case within the past 14 days; OR
  − Have fever and lower respiratory symptoms (cough, shortness of breath) and a negative rapid flu test
▪ Meteorological indicators are: average temperature in °C, Moisture %, wind km/h, days of rain and fog from 1st February to 1 April, 2020 (il Meteo, 2020).
▪ Interpersonal contact rates: a proxy here considers the density of cities (individual /km^2^) in 2019 (ISTAT, 2020).

### 1.3 Data analysis and procedure

This study analyses a database of *N*=55 Italian provincial capitals, considering variables in 2018-2019-2020 to explain the relationships between diffusion of COVID19, demographic, geographical and environmental variables. *Firstly*, preliminary analyses of variables are descriptive statistics based on mean, std. deviation, skewness and kurtosis to assess the normality of distributions and, if necessary to fix distributions of variables with a *log*-transformation.

Statistical analyses are also done categorizing Italian provincial capitals (*N*=55) in groups as follows:

− Hinterland cities
− Coastal cities

Categorization in:

− Windy cities
− Not windy cities

Categorization in:

− Cities of North Italy
− Cities of Central-South Italy

Categorization in:

− Cities with >100 days per year exceeding the limits set for PM_10_ or for ozone
− Cities with <100 days per year exceeding the limits set for PM_10_ or for ozone

Categorization in:

− Cities with ≤1000 inhabitant/km^2^
− Cities with > 1000 inhabitant/km^2^

Categorization in:

− Cities with ≤500 inhabitant/km^2^
− Cities with 500-1500 inhabitant/km^2^
− Cities with >1500 inhabitants/km^2^

*Secondly*, the bivariate and partial correlation verifies relationships (or associations) between variables understudy, and measures the degree of association. After that the null hypothesis (*H*_0_) and alternative hypothesis (*H*_1_) of the significance test for correlation is computed, considering two-tailed significance test.

*Thirdly*, the analysis considers the relation between independent and dependent variables. In particular, the dependent variable (number of infected people across Italian provincial capitals) is a linear function of a single explanatory variable given by total days of exceeding the limits set for PM_10_ across Italian province capitals. Dependent variables have in general *a lag of 1 years* in comparison with explanatory variables to consider temporal effects of air pollution predictor on environment and population in the presence of viral infectivity by COVID19 in specific cities of Italy.

The specification of the linear relationship is a *log-log* model is:

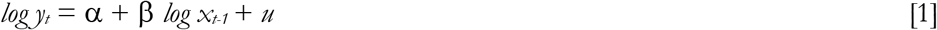

α is a constant; β= coefficient of regression; *u* = error term

*y* = dependent variable is number of infected individuals in cities

*x* = explanatory variable is a measure of air pollution, given by total days of exceeding the limits set for PM_10 or_ ozone in cities

This study extends the analysis with a multiple regression model to assess how different indicators can affect diffusion of COVID-19. The specification of the linear relationship is also a *log-log* model as follows:

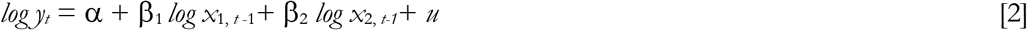

*y* = dependent variable is number of infected individuals in cities

*x* _*1*_ =explanatory variable is a measure of air pollution, given by total days of exceeding the limits set for PM_10 or_ ozone in cities

*x*_*2*_ = density of cities, inhabitants /km^2^

In addition, equation [2] is performed using data of infected at *t*=17^th^ March, 2020 in the starting phase of growth of the outbreak in Italy, and the at *t+16days*= 1^st^ April, 2020 in the phase of maturity of viral infectivity during lockdown and quarantine to assess the magnitude of two explanatory variables in the transmission dynamics of COVID-19. The estimation of equation [2] is also performed using hierarchical multiple regression, a variant of the basic multiple regression procedure that allows to specify a fixed order of entry for variables in order to control for the effects of covariates or to test the effects of certain predictors independent of the influence of others. The R^2^ changes are important to assess the predictive role of additional variables. The adjusted R-square and standard error of the estimate are useful as comparative measures to assess results between models. The *F*-test evaluates if the regression model is better than using only the mean of the dependent variable. If the *F* value is very small (e.g., 0.001), then the independent variables reliably predict the dependent variable.

Moreover, the linear relationship is also specified with a quadratic model as follows:

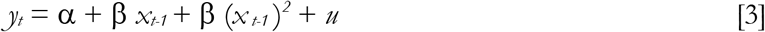

the goal is to apply an optimization approach, to calculate the minimum of equation [3] that suggests the maximum number of days in which cities can exceed the limits set for PM_10_. or ozone. Beyond this critical estimated limit, there are environmental inconsistencies of air pollution associated with meteorological conditions that can trigger a take-off of viral infectivity with damages for health of population and economic system (cf., Coccia, 2017c, 2017d). The max number of days in which cities can exceed the limit set for air pollution that minimizes the number of people infected, before the take-off of epidemic curve, can also suggest implications of proactive strategies and critical decision to cope with future epidemics similar to COVID-19 in society.

Finally, if *y*_t_ is number of infected individuals referred to a specific day, and equation [1] is calculated for each day changing dependent variable by using data of infected people in day *1*, day *2*, day *3*, …, day *n*, the variation of coefficient of regression *b*, such as during and after quarantine and lockdown can be used to assed the possible end of epidemic wave as follows:

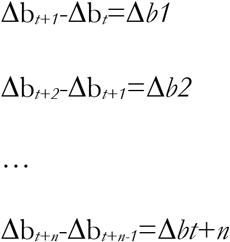

Average reduction is 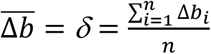

After that, decreasing b_*t*_ at *t* from day *1* to day *n* of the constant value 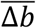, the i*-th* day when b_*t*_ is close to 0, it suggests the ending tail of epidemics. Ordinary Least Squares (OLS) method is applied for estimating the unknown parameters of relations in linear regression models [1-3]. Statistical analyses are performed with the Statistics Software SPSS® version 24.

## RESULTS

Descriptive statistics of variables in *log* scale, based on Italian province capitals (*N*=55), have normal distribution to apply appropriate parametric analyses.

Table 1 shows that hinterland cities have and average higher level of infected individuals than coastal cities. Hinterland cities have also a higher air pollution (average days per years) than coastal cities, in a context of meteorological factors of lower average temperature, lower average wind speed, lower rain days and lower level of moisture % than coastal cities.

**Table 1.** Descriptive statistics of Hinterland and Coastal Italian province capitals

|  | Days<br>exceeding<br>limits set<br>for PM<sub>10</sub><br>or ozone<br>2018 | Infected<br>17<sup>th</sup><br>March<br>2020 | Infected<br>1<sup>st</sup> April<br>2020 | Density<br>inhabitants/km<sup>2</sup><br>2019 | Temp °C<br>Feb-Mar<br>2020 | Moisture<br>%<br>Feb-Mar<br>2020 | Wind<br>km/h<br>Feb-Mer<br>2020 | Rain<br>Days<br>Feb-Mar<br>2020 | Fog<br>Days<br>Feb-Mar<br>2020 |
| --- | --- | --- | --- | --- | --- | --- | --- | --- | --- |
| *Hinterland cities N=45* |  |  |  |  |  |  |  |  |  |
| Mean | 80.40 | 497.00 | 1929.69 | 1480.11 | 9.11 | 68.31 | 8.02 | 4.81 | 4.14 |
| Std. Deviation | 41.66 | 767.19 | 2265.86 | 1524.25 | 2.20 | 7.68 | 3.69 | 2.38 | 3.13 |
| *Coastal cities N=10* |  |  |  |  |  |  |  |  |  |
| Mean | 59.40 | 171.30 | 715.80 | 1332.80 | 10.61 | 74.40 | 11.73 | 5.10 | 3.25 |
| Std. Deviation | 38.61 | 164.96 | 522.67 | 2463.04 | 2.20 | 7.38 | 2.60 | 2.71 | 3.68 |

Table 2 shows that cities with low intensity of wind speed (7.3km/h) have and average higher level of infected individuals than windy cities (average of 12.77km/h). Cities with lower intensity of wind speed have also a higher level of air pollution (average days per years), in a meteorological context of lower average temperature, lower rain days, lower level of moisture % and a higher average days of fog.

**Table 2.** Descriptive statistics of windy and not windy of Italian province capitals

|  | Days<br>exceeding<br>limits set<br>for PM<sub>10</sub><br>or ozone<br>2018 | Infected<br>17<sup>th</sup><br>March<br>2020 | Infected<br>1<sup>st</sup> April<br>2020 | Density<br>inhabitants/km<sup>2</sup><br>2019 | Temp °C<br>Feb-Mar<br>2020 | Moisture<br>%<br>Feb-Mar<br>2020 | Wind<br>km/h<br>Feb-Mer<br>2020 | Rain<br>Days<br>Feb-Mar<br>2020 | Fog<br>Days<br>Feb-Mar<br>2020 |
| --- | --- | --- | --- | --- | --- | --- | --- | --- | --- |
| *Low windy cities N=41* |  |  |  |  |  |  |  |  |  |
| Mean | 84.32 | 536.20 | 2036.15 | 1517.41 | 9.05 | 68.23 | 7.30 | 4.56 | 4.18 |
| Std. Deviation | 43.31 | 792.84 | 2333.72 | 1569.70 | 2.12 | 7.50 | 2.77 | 2.33 | 2.94 |
| *High windy cities N=14* |  |  |  |  |  |  |  |  |  |
| Mean | 53.93 | 149.57 | 750.86 | 1265.64 | 10.36 | 72.89 | 12.77 | 5.75 | 3.39 |
| Std. Deviation | 25.87 | 153.55 | 640.02 | 2108.31 | 2.43 | 8.37 | 3.46 | 2.56 | 4.00 |

Table 3 shows that cities in the central and southern part of Italy have, during the COVID-19 outbreak, a lower number of infected than cities in North Italy. This result is in an environment with lower air pollution (average days per years), higher average temperature, higher average wind speed, higher rain days and lower level of moisture %.

**Table 3.** Descriptive statistics of Northern and Central-Southern Italian province capitals

|  | Days<br>exceeding<br>limits set<br>for PM<sub>10</sub><br>or ozone<br>2018 | Infected<br>17<sup>th</sup><br>March<br>2020 | Infected<br>1<sup>st</sup> April<br>2020 | Density<br>inhabitants/km<sup>2</sup><br>2019 | Temp °C<br>Feb-Mar<br>2020 | Moisture<br>%<br>Feb-Mar<br>2020 | Wind<br>km/h<br>Feb-Mer<br>2020 | Rain<br>Days<br>Feb-Mar<br>2020 | Fog<br>Days<br>Feb-Mar<br>2020 |
| --- | --- | --- | --- | --- | --- | --- | --- | --- | --- |
| *Norther cities N=45* |  |  |  |  |  |  |  |  |  |
| Mean | 80.51 | 515.60 | 1968.42 | 1448.00 | 9.05 | 69.40 | 7.89 | 4.80 | 4.31 |
| Std. Deviation | 42.67 | 759.18 | 2230.43 | 1538.10 | 1.97 | 7.61 | 3.15 | 2.42 | 3.06 |
| *Central-Southern cities*<br>*N=10* |  |  |  |  |  |  |  |  |  |
| Mean | 58.90 | 87.60 | 541.50 | 1477.30 | 10.88 | 69.50 | 12.31 | 5.15 | 2.50 |
| Std. Deviation | 32.36 | 129.98 | 735.21 | 2424.50 | 2.92 | 9.64 | 4.44 | 2.52 | 3.65 |

Table 4 confirms previous results considering cities with >100days exceeding limits set for PM_10_ or ozone: they have, *versus* cities with less than 100 days, a very high level of infected individuals, in an environment of higher average density of population, lower average intensity of wind speed, lower average temperature with higher average moisture % and days of fog.

**Table 4.**
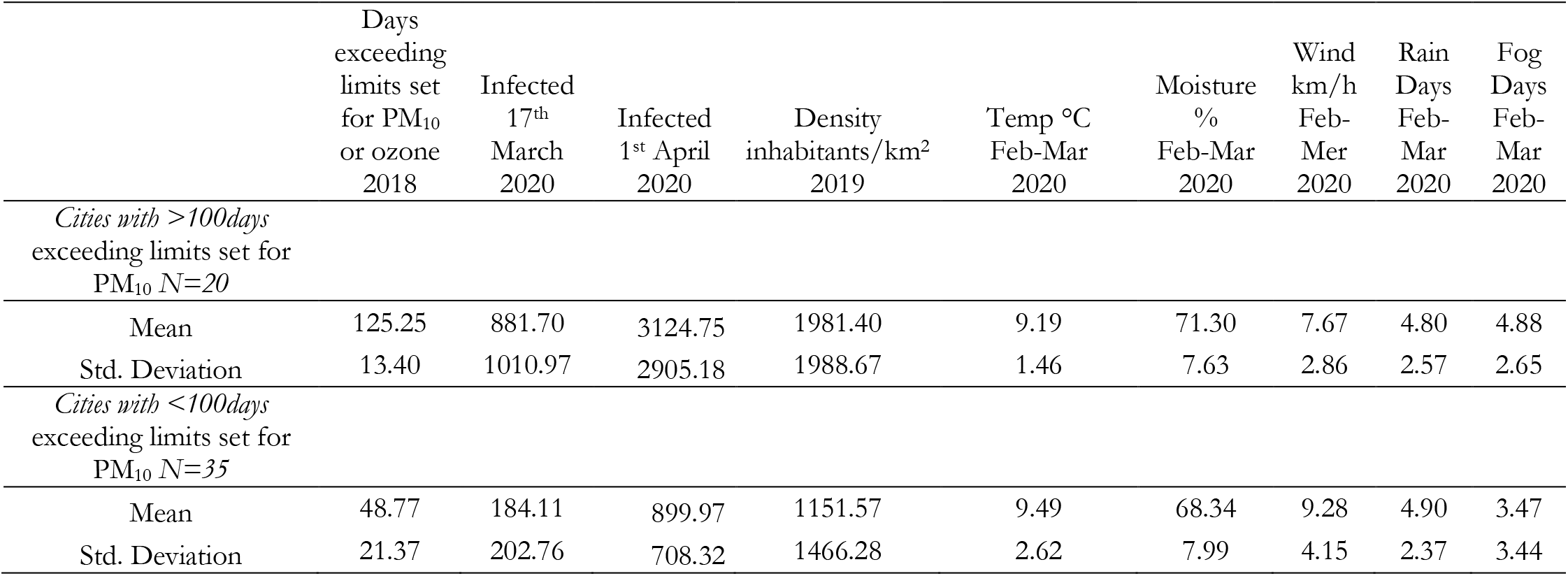
Descriptive statistics of Italian provincial capitals according to days exceeding the limits set for PM_10_

Tables 5-6 show results considering categorization of cities per density of population/km^2.^. Results reveal that average number of infected individuals increases with average density of people/km^2^, but with an arithmetic growth, in comparison to geometric growth of number of infected individuals with other categorizations of cities. These findings suggest that density of population per km^2^ is important for transmission dynamics but other factors may support acceleration of viral infectivity by COVID-19 rather than high probability of interpersonal contacts in cities. In short, results suggest that among Italian province capitals:

- Number of infected people is HIGHER in: Cities with >100days exceeding limits set for PM_10_ or ozone, located in hinterland zones having a low average intensity of wind speed and lower temperature in °C.

**Table 5.** Descriptive statistics of Italian provincial capitals according to density per km^2^ (2 categories)

|  | Days<br>exceeding<br>limits set<br>for PM<sub>10</sub><br>or ozone<br>2018 | Infected<br>17<sup>th</sup><br>March<br>2020 | Infected<br>1<sup>st</sup> April<br>2020 | Density<br>inhabitants/km<sup>2</sup><br>2019 | Temp °C<br>Feb-Mar<br>2020 | Moisture<br>%<br>Feb-Mar<br>2020 | Wind<br>km/h<br>Feb-Mer<br>2020 | Rain<br>Days<br>Feb-Mar<br>2020 | Fog<br>Days<br>Feb-Mar<br>2020 |
| --- | --- | --- | --- | --- | --- | --- | --- | --- | --- |
| *Cities with ≤1000*<br>*inhabitant/km<sup>2</sup>*<br>*N=30* |  |  |  |  |  |  |  |  |  |
| Mean | 64.37 | 248.37 | 960.97 | 510.77 | 10.01 | 69.61 | 9.28 | 4.08 | 3.75 |
| Std. Deviation | 39.25 | 386.95 | 951.26 | 282.11 | 1.95 | 10.30 | 4.41 | 2.37 | 3.40 |
| *Cities with >1000*<br>*inhabitant/km<sup>2</sup>*<br>*N=25* |  |  |  |  |  |  |  |  |  |
| Mean | 91.24 | 665.08 | 2606.60 | 2584.40 | 8.63 | 69.19 | 7.99 | 5.80 | 4.26 |
| Std. Deviation | 40.24 | 919.70 | 2717.57 | 2000.63 | 2.40 | 3.59 | 2.79 | 2.17 | 3.03 |

**Table 6.** Descriptive statistics of Italian provincial capitals according to density per km^2^ (2 categories)

|  | Days<br>exceeding<br>limits set<br>for PM<sub>10</sub><br>or ozone<br>2018 | Infected<br>17<sup>th</sup><br>March<br>2020 | Infected<br>1<sup>st</sup> April<br>2020 | Density<br>inhabitants/km<sup>2</sup><br>2019 | Temp °C<br>Feb-Mar<br>2020 | Moisture<br>%<br>Feb-Mar<br>2020 | Wind<br>km/h<br>Feb-Mer<br>2020 | Rain<br>Days<br>Feb-Mar<br>2020 | Fog<br>Days<br>Feb-Mar<br>2020 |
| --- | --- | --- | --- | --- | --- | --- | --- | --- | --- |
| *Cities with <1000*<br>*inhabitant/km<sup>2</sup>*<br>*N=17* |  |  |  |  |  |  |  |  |  |
| Mean | 52.82 | 116.12 | 567.12 | 312.76 | 9.88 | 71.12 | 9.52 | 4.44 | 4.41 |
| Std. Deviation | 36.87 | 128.13 | 466.49 | 161.34 | 2.12 | 8.91 | 5.73 | 2.79 | 3.79 |
| *Cities with 500-1000*<br>*inhabitant/km<sup>2</sup>*<br>*N=22* |  |  |  |  |  |  |  |  |  |
| Mean | 84.32 | 430.91 | 1519.50 | 951.32 | 9.04 | 68.50 | 8.37 | 4.34 | 3.75 |
| Std. Deviation | 37.28 | 476.29 | 1018.07 | 277.77 | 2.65 | 9.17 | 2.33 | 2.06 | 2.99 |
| *Cities with >1000*<br>*inhabitant/km<sup>2</sup>*<br>*N=16* |  |  |  |  |  |  |  |  |  |
| Mean | 91.19 | 789.00 | 3182.75 | 3355.44 | 9.33 | 68.86 | 8.26 | 6.03 | 3.84 |
| Std. Deviation | 43.29 | 1103.03 | 3239.96 | 2151.27 | 1.81 | 4.32 | 2.79 | 2.19 | 3.03 |

Table 7 shows association between variables on 17^th^ March and 1^st^ April, 2020: a correlation higher than 62% (*p*-value<.001) is between air pollution and infected individuals, a lower coefficient of correlation is between density of population and infected individuals (*r*=48-55%, *p*-value<.001). Results also show a negative correlation between number of infected individuals and intensity of wind speed among cities (*r*= –28 to –38%, *p*-value <0.05): this effect is due to the role of wind speed that cleans air from pollutants that are associated with transmission dynamics of viral infectivity.

**Table 7.**
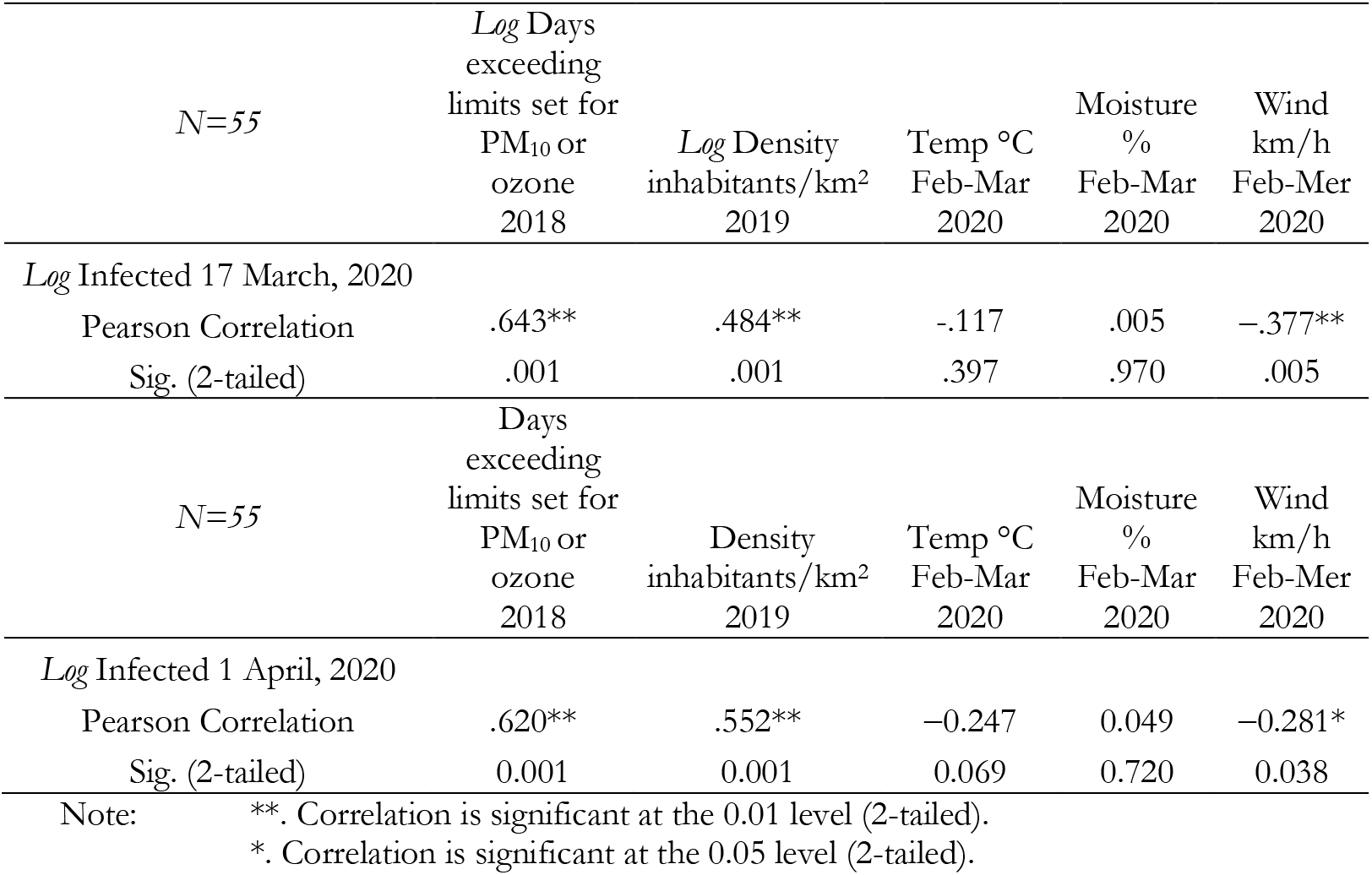
Correlation

Table 8 confirms the high correlation between air pollution and infected individuals on 17^th^ March and 1 April, 2020, controlling meteorological factors of cities under study (*r*>60%, *p*-value<.001).

**Table 8.** Partial Correlation

| *Control Variables*<br>Temp °C<br>Moisture %<br>Wind km/h<br>Feb-Mar 2020 | *Pearson Correlation* | *Log*<br>Infected<br>17 March,<br>2020 | *Log*<br>Infected<br>1 April,<br>2020 |
| --- | --- | --- | --- |
|  | *Log Days exceeding limits*<br>set for PM<sub>10</sub> or ozone<br>2018 | 0.607 | .602 |
|  | Sig. (2-tailed) | .001 | .001 |
|  | N | 50 | 50 |

Partial correlation in table 9 suggests that controlling density of population on 17^th^ march and 1^st^ April 2020, number of infected people is associated with air pollution (*r*≥50%, *p*-value<.001), whereas, controlling air pollution the correlation between density of population in cities and infected individuals is lower (*r*=27-38%, *p*-value<.001). The reduction of *r* between infected individuals and air pollution from 17^th^ March to 1^st^ April, and the increase of the association between infected people and density of people in cities over the same time period, controlling mutual variables, suggests that that air pollution in cities seems to be a more important factor in the initial phase of transmission dynamics of COVID-19 (i.e., 17^th^ March, 2020). In the phase of the maturity of transmission dynamics (1^st^ April, 2020), with lockdown that reduces air pollution, the role of air pollution reduces intensity whereas human-to-human transmission increases.

**Table 9.** Partial Correlation

| *Control Variables*<br>*Log* Density<br>inhabitants/km<sup>2</sup><br>2019 | *Pearson Correlation* | *Log*<br>Infected<br>17 March,<br>2020 | *Log*<br>Infected<br>1 April,<br>2020 |
| --- | --- | --- | --- |
|  | *Log Days exceeding limits*<br>set for PM<sub>10</sub> or ozone<br>2018 | 0.542 | .496 |
|  | Sig. (2-tailed) | .001 | .001 |
|  | N | 52 | 52 |
| *Control Variables*<br>*Log* Days exceeding limits<br>set for PM<sub>10</sub> or ozone<br>2018 | *Pearson Correlation* | *Log*<br>Infected<br>17 March,<br>2020 | *Log*<br>Infected<br>1 April,<br>2020 |
|  | *Log* Density<br>inhabitants/km<sup>2</sup><br>2019 | 0.279 | .385 |
|  | Sig. (2-tailed) | .041 | .004 |
|  | N | 50 | 50 |

These findings are confirmed with hierarchical regression that also reveals how air pollution in cities seems to be a driving factor of transmission dynamics in the growing phase of CIVID-19 (17^th^ March, 2020). In the phase of the maturity of transmission dynamics (1^st^ April, 2020), the determinant of air pollution is important to support infected population but reduces intensity, whereas the factor of human-to-human transmission increases, *ceteris paribus* (Table 10). <u>This result reveals that transmissions dynamics of COVID-19 is due to human-to-human transmission but the factor of air pollution-to-human transmission of viral infectivity supports a substantial growth.</u>

**Table 10.**
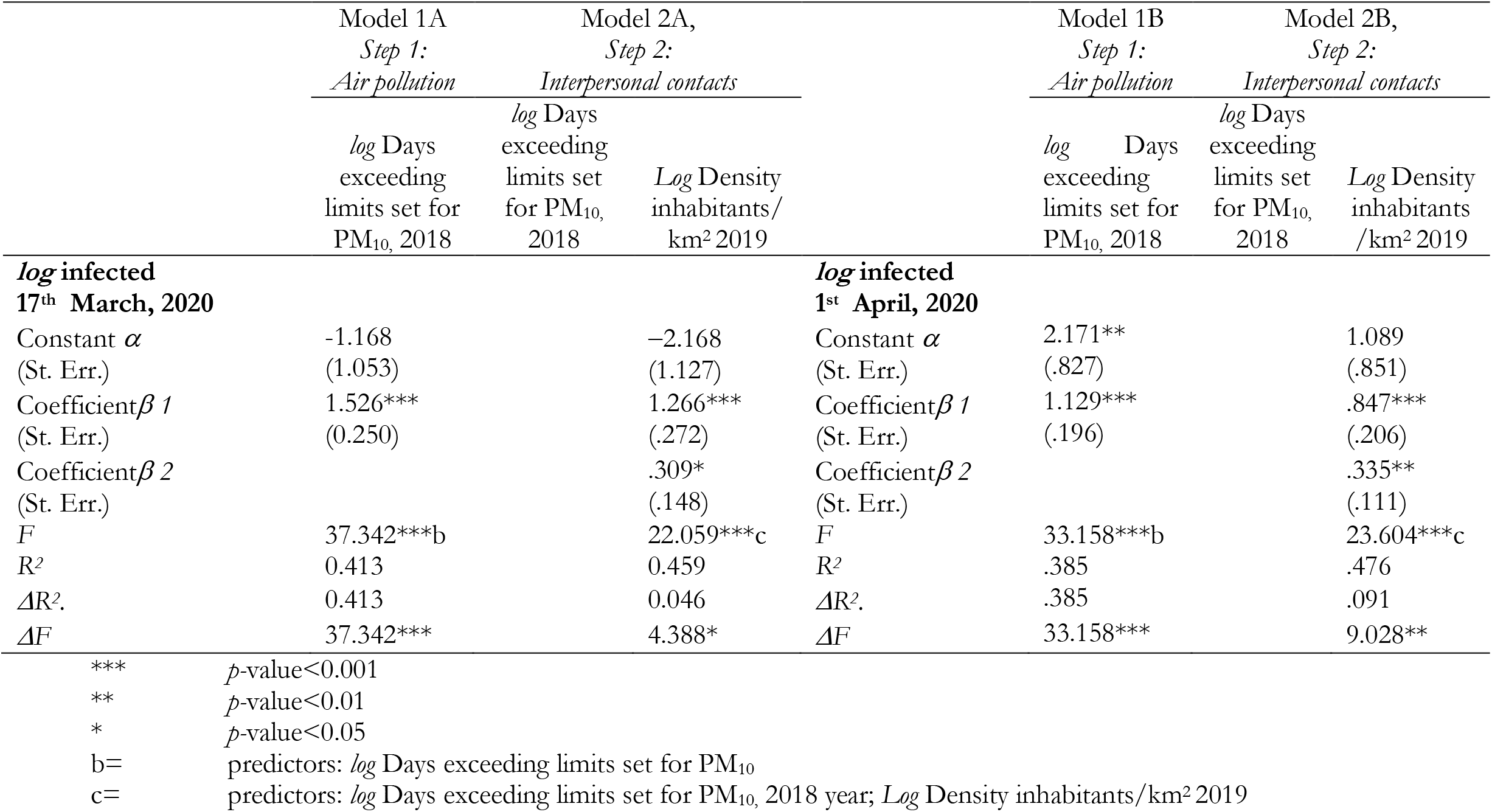
Parametric estimates of the relationship of *Log* Infected 17 March and 1 April on *Log* Days exceeding limits set for PM_10_ and *Log* Density inhabitants/km^2^ 2019 (*hierarchical regression*)

Table 11 shows results of the transmission dynamics of COVID-19 considering the interpersonal contacts, measured with density of population in cities understudy. In short, results suggest that density of population explains the number of infected individuals, increasing the probability of human-to-human transmission. However, if we decompose the sample to consider the cities with ≤100 days exceeding limits set for PM_10_ or ozone and with >100 days exceeding limits set for PM_10_ or ozone, then the expected increase of number of infected individuals is higher in cities having more than 100 days exceeding limits set for PM_10_ or ozone. In particular,

○ Cities with ≤100 days exceeding limits set for PM_10_, an increase of 1% in density of population, it increases the expected number of infected by about 0.30%
○ Cities with >100 days exceeding limits set for PM_10_, an increase of 1% in density of population, it increases the expected number of infected by about 1.43%!

**Table 11.** Parametric estimates of the relationship of *Log* Infected 1^st^ April,2020 on *Log* Density inhabitants/km^2^ 2019, considering the groups of cities *with* days exceeding limits set for PM_10_ or ozone

| ↓DEPENDENT VARIABLE | Model cities with <100 days<br>exceeding limits set for PM<sub>10</sub><br>or ozone, 2018 | ↓DEPENDENT VARIABLE | Model cities with >100 days<br>exceeding limits set for PM<sub>10</sub><br>or ozone, 2018 |
| --- | --- | --- | --- |
|  | *Log* Density inhabitants/km<sup>2</sup><br>2019 |  | *Log* Density inhabitants/km<sup>2</sup><br>2019 |
| ***log* infected**<br>**1 April, 2020** |  | ***log* infected**<br>**1 April, 2020** |  |
| Constant $\alpha$ | 4.501 | Constant $\alpha$ | 1.425 |
| (St. Err.) | (.801) | (St. Err.) | (1.624) |
| Coefficient $\beta$ 1 | 0.303* | Coefficient $\beta$ 1 | 0.856*** |
| (St. Err.) | (0.122) | (St. Err.) | (0.223) |
| R<sup>2</sup> (St. Err. of Estimate) | 0.158 (.828) | R<sup>2</sup> (St. Err. of Estimate) | 0.450(.803) |
| F | 6.207* | F | 14.714*** |

Note: Explanatory variable: *Log* Density inhabitants/km<sup>2</sup> in 2019
\*\*\* $p$ -value<0.001
\* $p$ -value<0.05

The statistical output of table 11 is schematically summarized as follows:

|  | Cities with ≤100 days exceeding limits set for PM<sub>10</sub> | Cities with >100 days exceeding limits set for PM<sub>10</sub> |
| --- | --- | --- |
| Density of population | 0.30 ($p<0.05$) | 1.43 ($p<0.001$) |
| F | 6.207 ($p<0.05$) | 14.714 ($p<0.001$) |
| R<sup>2</sup> | 15.8% | 45% |

Figure 1 confirms, *ictu oculi*, that the coefficient of regression in cities with >100 days exceeding limits set for PM_10_ is much bigger than the coefficient in cities with ≤100 days exceeding limits set for PM_10_, <u>suggesting that air pollution- to-human transmission is definitely important to explain the transmission dynamics of COVID-19.</u> The policy implications here are clear: COVID-19 has reduced transmission dynamics on population in the presence of lower level of air pollution and specific environments with lower intensity of wind speed. Hence, the effect of accelerated transmission dynamics of COVID-19 cannot be explained without accounting for the level of air pollution and geo-environmental conditions of the cities.

**Figure 1:**
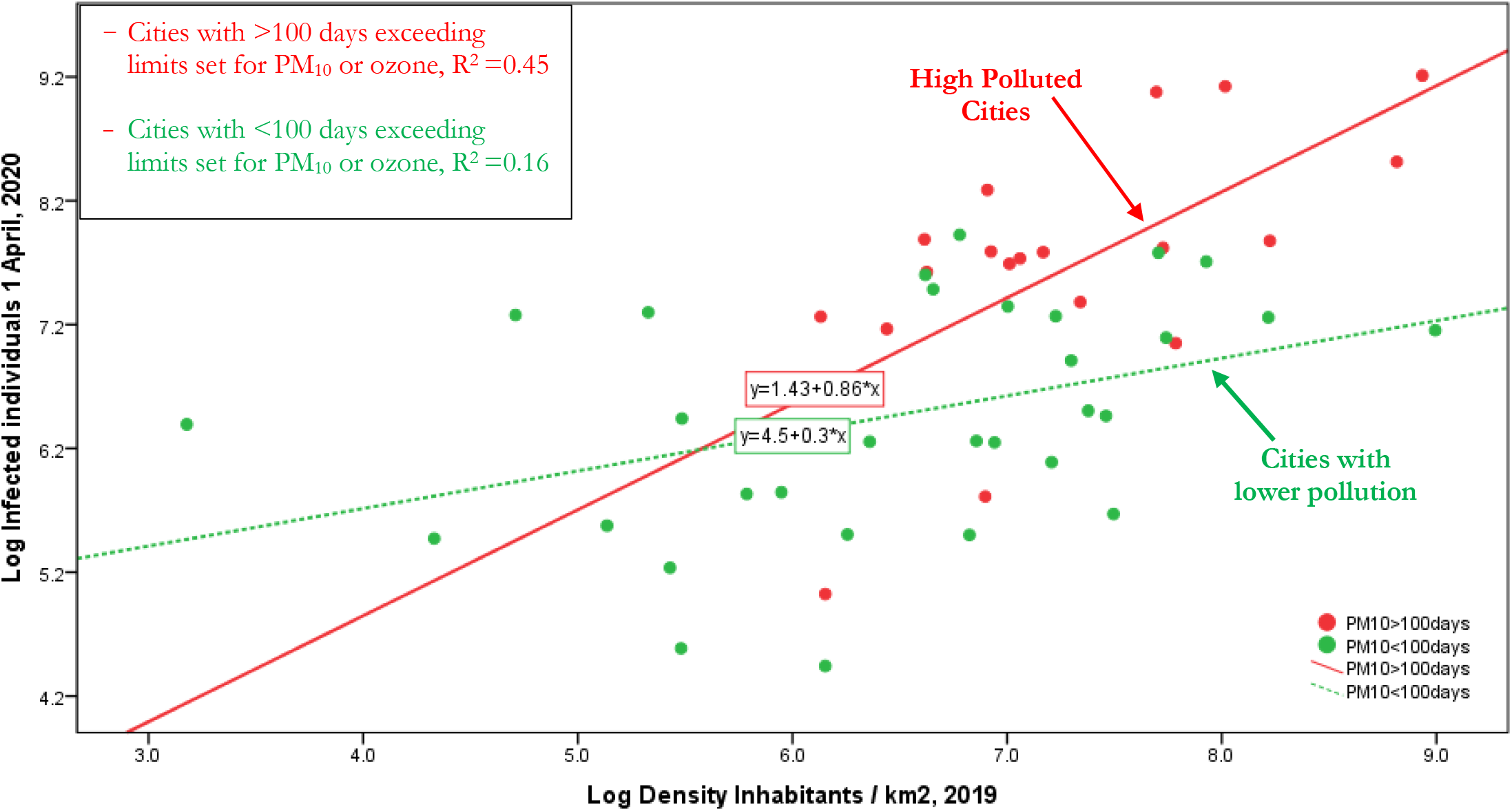
Regression line of *Log* Infected 1 April,2020 on *Log* Density inhabitants/km^2^ 2019, considering the groups of cities *with* days exceeding limits set for PM_10_ or ozone <, or ≥100 days. Note: *This result reveals that transmissions dynamics of COVID-19 is due to human-to-human transmission* (density of population) *but in polluting cities the accelerated diffusion is due to air pollution-to-human transmission of viral infectivity*

A main question for environmental policy is: What is the maximum number of days in which cities can exceed the limits set for PM_10_ or ozone per year, before that the combination between air pollution and meteorological condition triggers a take-off of viral infectivity (epidemic diffusion) with damages for health of population and economy in society?

The function based on table 12 is:

*y* = 1438.808–35.322 *x*+0.393 *x*^2^

*y* = number of infected individuals 1^st^ April, 2020

*x*= days exceeding limits of PM_10_ or ozone in Italian provincial capitals

The minimization is performed imposing first derivative equal to zero.

**Table 12.**
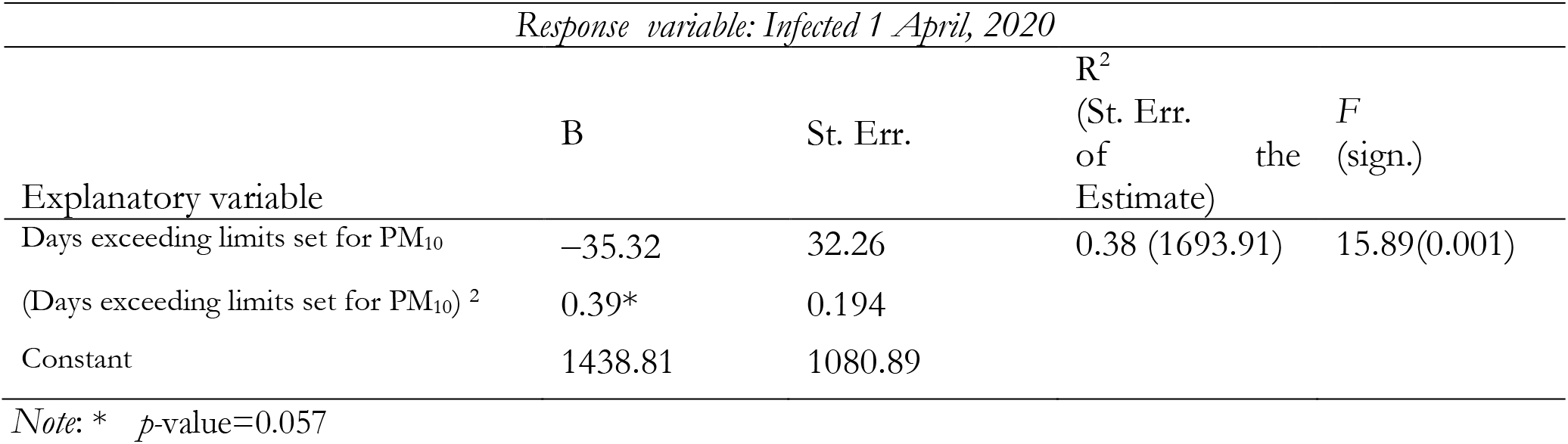
Parametric estimates of the relationship of Infected 1^st^ April, 2020 on days exceeding limits set for PM10 (*simple regression analysis, quadratic model*)

D*y / x* =*y’* =*–*35.322+0.786*x* = *0*

*x*=35.32/0.786= 44.94 ∼ 45 *days* exceeding limits of PM_10_ or ozone in Italian provincial capitals (cf., Figure 2).

**Figure 2.**
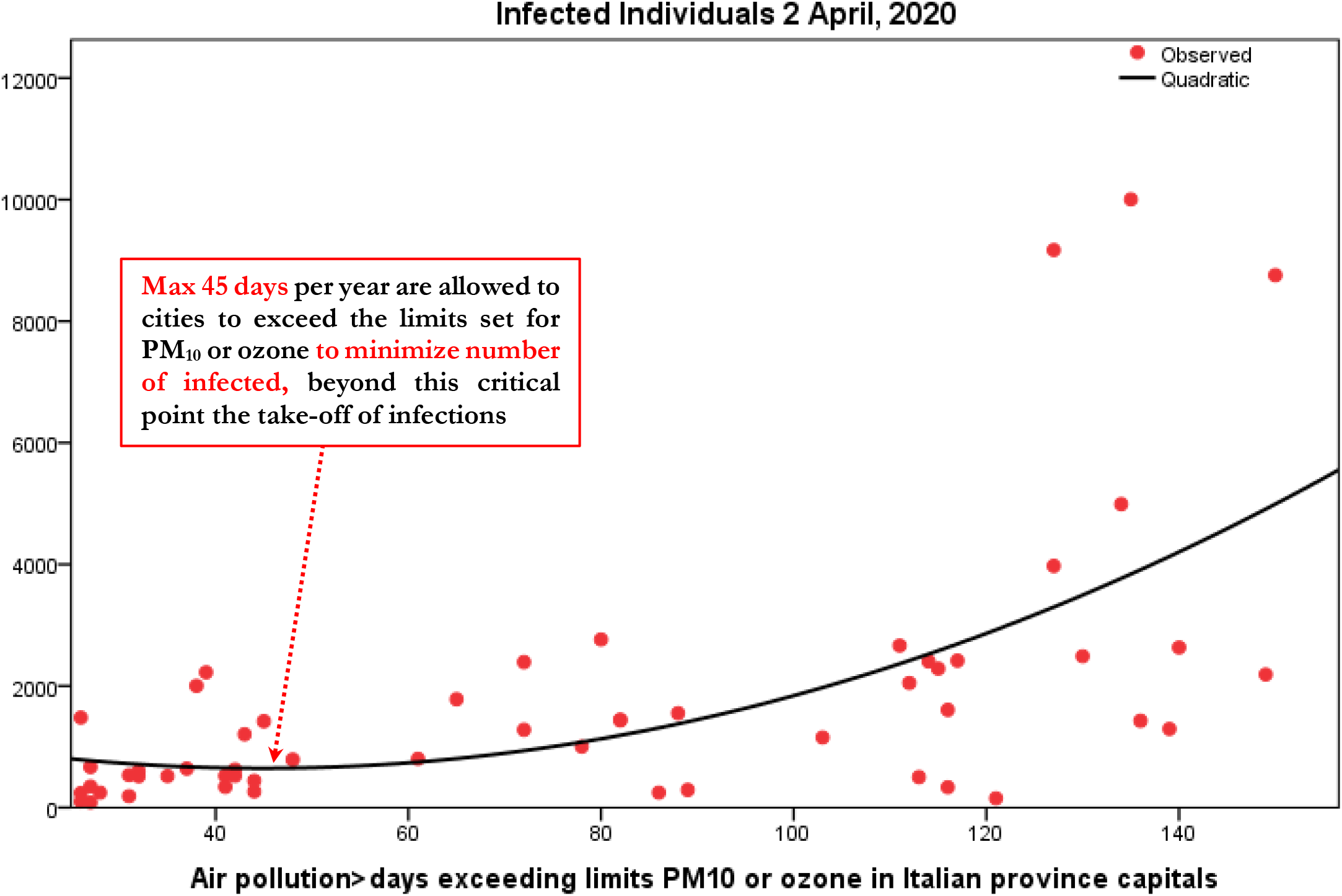
Estimated relationship of number of infected on days exceeding limits of PM_10_ in Italian provincial capitals (*Quadratic model*)

This finding suggests that the max number of days in which Italian provincial capitals can exceed per year the limits set for PM_10_ (particulate matter 10 micrometers or less in diameter) or for ozone, considering the meteorological condition is about 45 days. Beyond this critical point, the analytical and geometrical output suggests that environmental inconsistencies because of the combination between air pollution and meteorological conditions trigger a take-off of viral infectivity (epidemic diffusion) with damages for health of population and economy in society.

Finally, the reduction of unstandardized coefficient of regression in table 13, from 17^th^ March to 1^st^ April, suggests a declining trend of COVID-19 viral infectivity over time and space. The question is: *how many days are necessary to stop the epidemic, ceteris paribus* (*quarantine and lockdown*)*?*

**Table 13.** Coefficient of regression of linear model per day based on equation [1] and daily change

| day | Unstandardized<br>coefficient B | Standard<br>Error | Sign. | R<sup>2</sup> | $\Delta=B_t-B(t-1)$ |
| --- | --- | --- | --- | --- | --- |
| 17<sup>th</sup> March | 1.526 | 0.250 | 0.001 | 0.400 |  |
| 19 | 1.362 | 0.238 | 0.001 | 0.370 | -0.1640 |
| 21 | 1.360 | 0.234 | 0.001 | 0.377 | -0.0020 |
| 22 | 1.340 | 0.232 | 0.001 | 0.375 | -0.0200 |
| 23 | 1.322 | 0.231 | 0.001 | 0.370 | -0.0180 |
| 24 | 1.320 | 0.228 | 0.001 | 0.376 | -0.0020 |
| 225 | 1.285 | 0.227 | 0.001 | 0.364 | -0.0350 |
| 26 | 1.287 | 0.223 | 0.001 | 0.375 | 0.0020 |
| 27 | 1.291 | 0.221 | 0.001 | 0.380 | 0.0040 |
| 28 | 1.289 | 0.202 | 0.001 | 0.424 | -0.0020 |
| 29 | 1.246 | 0.216 | 0.001 | 0.375 | -0.0430 |
| 30 | 1.236 | 0.209 | 0.001 | 0.386 | -0.0100 |
| 31 | 1.143 | 0.198 | 0.001 | 0.375 | -0.0930 |
| 1<sup>st</sup> April | 1.162 | 0.201 | 0.001 | 0.376 | 0.0190 |
| 2<sup>nd</sup> April | 1.129 | 0.196 | 0.001 | 0.373 | -0.0330 |
| Arithmetic mean $\Delta = \delta =$ | | | | | -0.0284 |

Note: Coefficients of regression at day $t$ is calculated using data at $t-1$

Let, the average reduction of the coefficient of regression at *t*=2^nd^ April, 2020 equal to δ=–0.0284, let B at 2^nd^ April, 2020 (on data updated at 1st April, 2020), B=1.129, the days necessary to reduce B close to 0 (*zero*), with a constant reduction per day equal to δ, calculated as explained in methods of this study here, is about 40 days (i.e., by 15 May, 2020), when B may be lower than 0.05, fixed current conditions of quarantine and lockdown.

## DISCUSSION

Considering the results just mentioned, the fundamental questions are:

*Why did this virus spread so rapidly in Italy?*

*How is the link between geographical and environmental factors and accelerate diffusion of COVID-19 in specific regions?*

Figure 3 show COVID-19 outbreak in North Italy with number of infected and days exceeding the limits set for PM_10_ or for ozone. Statistical analyses for *N*=55 Italian provincial capitals confirm the significant association between high diffusion of viral infectivity and air pollution. Studies show that the diffusion of viral infectivity depends on the interplay between host factors and the environment (Neu and Mainou, 2020). In this context, it is critical to understand how air quality can affect viral dissemination at national and global level (Das and Horton, 2017). Many ecological studies have examined the association between the incidence of invasive pneumococcal disease and respiratory virus circulation and various climatic factors (McCullers, 2006; Jansen et al., 2008). These studies show that in temperate climates, the epidemiology of invasive pneumococcal disease has a peak incidence in winter months (Dowell et al., 2003; Kim et al., 1996; Talbot et al. 2005). Brunekreef and Holgate (2002) argue that, in addition to climate factors, the health effects of air pollution have been subject to intense investigations in recent years. Air pollution is ubiquitous in manifold urban areas worldwide of developed and developing nations. Air pollution has gaseous components and particulate matter (PM). The former includes ozone (O_3_), volatile organic compounds (VOCs), carbon monoxide (CO) and nitrogen oxides (NO_x_) that generate inflammatory stimuli on the respiratory tract (Glencross et al., 2020). Of these pollutants, PM has a complex composition that includes metals, elemental carbon and organic carbon (both in hydrocarbons and peptides), sulphates and nitrates, etc. (Ghio et al., 2012; Wooding et al., 2019).

**Figure 3.**
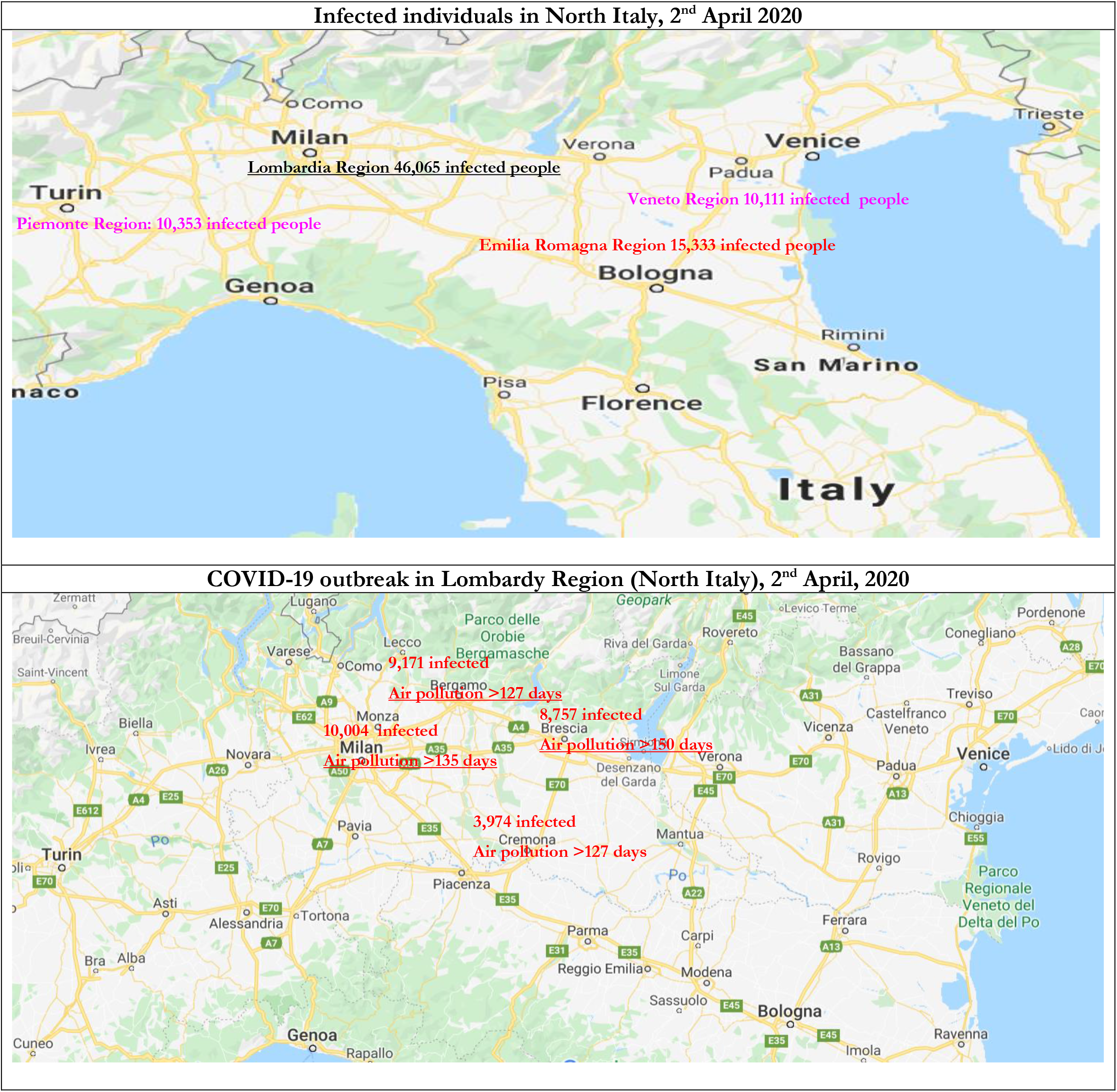
COVID-19 Outbreak (number of infected individual on 2^nd^ April, 2020) and days exceeding the limits set for PM_10_ or ozone

Advanced countries, such as in Europe, have more and more smog because of an unexpected temperature inversion, which trap emissions from the city’s coal-burning heating stoves and diesel powered buses near ground-level in winter. The ambient pollution mixes with moisture in the air to form a thick, foul-smelling fog that affect the health of people in the city (Wang et al., 2016; Bell et al., 2004). The exposure to pollutants, such as airborne particulate matter and ozone, generates respiratory and cardiovascular diseases with increases in mortality and hospital admissions (cf., Langrish and Mills, 2014). Wei et al. (2020) analyze the effect of heavy aerosol pollution in northern China–characterized by long-duration, high PM_2.5_ concentrations and wide geographical coverage– that impacts on environmental ecology, climate change and public health (cf., Liu et al., 2017, 2018; Jin et al., 2017). The biological components of air pollutants and bio aerosols also include bacteria, viruses, pollens, fungi, and animal/plant fragments (Després et al., 2012; Fröhlich-Nowoisky et al., 2016; Smets et al., 2016). Studies show that during heavy aerosol pollution in Beijing (China), 50%-70% of bacterial aerosols are in sub micrometer particles, 0.56-1 mm (Zhang et al., 2019; cf., Zhang et al., 2016). As bacteria size typically ranges from 0.5 to 2.0 mm (Després et al., 2012), they can form clumps or attach to particles and transport regionally between terrestrial, aquatic, atmospheric and artificial ecosystems (Smets et al., 2016). Moreover, because of regional bio aerosol transportation, harmful microbial components, bacterial aerosols have dangerous implications on human health and also plantation (cf., Van Leuken et al., 2016). Harmful bio aerosol components–including pathogens, antibiotic-resistant bacteria, and endotoxins–can cause severe respiratory and cardiovascular diseases in society (Charmi et al., 2018). In fact, the concentration of microbes, pathogens and toxic components significantly increases during polluted days, compared to no polluted days (Liu et al., 2018). In addition, airborne bacterial community structure and concentration varies with pollutant concentration, which may be related to bacterial sources and multiplication in the air (Zhang et al., 2019). Studies also indicate that microbial community composition, concentration, and bioactivity are significantly affected by particle concentration (Liu et al., 2018). To put it differently, the atmospheric particulate matter harbors more microbes during polluted days than sunny or clean days (Wei et al., 2016). <u>These studies can explain one of the driving factors of higher viral infectivity of COVID-19 in the industrialized regions of Nord Italy, rather than other part of Italy</u> (*Tables 1-6*). In fact, viable bio aerosol particles and high microbial concentration in particulate matter play their non-negligible role during air pollution and transmission of viral infectivity (Zhang et al., 2019). For instance, airborne bacteria in PM_2.5_ from the Beijing-Tianjin-Hebei regions in China revealed that air pollutants are main factors in shaping bacterial community structure (Gao et al., 2017). Xie et al. (2018) indicate that total bacteria concentration is higher in moderately polluted air than in clean or heavily polluted air. Liu et al. (2018) show that bacterial concentration is low in moderately or heavily pollution in PM_2.5_ and PM_10_, whereas the pathogenic bacteria concentration is very high in heavy and moderate pollution. Sun et al. (2018) study bacterial community during low and high particulate matter (PM) pollution and find out that predominant species varied with PM concentration. In general, bio aerosol concentrations are influenced by complex factors, such as emission sources, terrain, meteorological conditions and other climate factors (Zhai et al., 2018). Wei et al. (2020) also investigate the differences between inland and coastal cities in China (Jinan and Weihai, respectively) to explain the influence of topography, meteorological conditions and geophysical factors on bio aerosol. Results suggest that from clean days to severely polluted days, bacterial community structure is influenced by bacterial adaptation to pollutants, chemical composition of pollutants and meteorological conditions (cf., Sun et al., 2018). Moreover, certain bacteria from Proteobacteria and Deinococcus-Thermus have high tolerance towards environmental stresses and can adapt to extreme environments. As a matter of fact, bacilli can survive to harsh environments by forming spores. Moreover, certain bacteria with protective mechanisms can survive in highly polluted environments, while other bacteria cannot withstand such extreme conditions. In particular, bacteria in the atmosphere to survive must withstand and adapt to ultraviolet exposure, reduced nutrient availability, desiccation, extreme temperatures and other factors. In addition, in the presence of accumulated airborne pollutants, more microorganisms might be attached to particulate matter. <u>Thus, in heavy or severe air pollution, highly toxic pollutants in PM_2.5_ and PM_10_ may inhibit microbial growth.</u> Numerous studies also indicate the role of meteorological conditions in pollution development that creates appropriate conditions for microbial community structure and abundance, and viral infectivity (Jones and Harrison, 2004). Zhong et al. (2018) argue that static meteorological conditions may explain the increase of PM_2.5_. In general, bacterial communities during aerosol pollution are influenced by bacterial adaptive mechanisms, particle composition, and meteorological conditions. The particles could also act as carriers, which have complex adsorption and toxicity effects on bacteria (Wei et al., 2020). Certain particle components are also available as nutrition for bacteria and the toxic effect dominates in heavy pollution. The differences in bacterial adaptability towards airborne pollutants cause bacterial survival or death for different species. Groulx et al. (2018) argue that microorganisms, such as bacteria and fungi in addition to other biological matter like endotoxins and spores comingle with particulate matter (PM) air pollutants. Hence, microorganisms may be influenced by interactions with ambient particles leading to the inhibition or enhancement of viability and environmental stability (e.g., tolerance to variation in seasonality, temperature, humidity, etc.). Moreover, <u>Groulx et al. (2018) claim that in the case of microbial agents of communicable disease, such as viruses, the potential for interactions with pollution may have public health implications.</u> Groulx et al. (2018, p. 1106) describe an experimental platform to investigate the implications of viral infectivity changes:

> Preliminary evidence suggests that the interactions between airborne viruses and airborne fine particulate matter influence viral stability and infectivity ….. The development of a platform to study interactions between artificial bio aerosols and concentrated ambient particles provides an opportunity to investigate the direction, magnitude and mechanistic basis of these effects, and to study their health implications.… The interactions of PM2.5 with Φ6 bacteriophages decreased viral infectivity compared to treatment with HEPA^2^-filtered air alone; By contrast, ΦX174, a non-enveloped virus, displayed increased infectivity when treated with PM2.5 particles relative to controls treated only with HEPA-filtered air.

Thus, the variation in bacterial community structure is related to different pollution intensities. Wei et al. (2020) show that Staphylococcus increased with PM_2.5_ and became the most abundant bacteria in moderate pollution. In heavy or severe pollution, bacteria, which are adaptable to harsh environments, increase. In moderate pollution, the PM_2.5_ might harbor abundant bacteria, especially genera containing opportunistic pathogens. Therefore, effective measures should control health risks caused by bio aerosols during air pollution, especially for immunocompromised, elderly and other fragile individuals. <u>This may explain the high mortality of certain individuals having previous pathologies because of COVID-19 in Italy that has the mortality rate (the percentage of deaths compared to the total of those</u> who tested positive for COVID-19) of about 80% in individuals aged > 70 years with comorbidities as of April 1^st^, <u>2020 (Istituto Superiore Sanità, 2020; cf., WHO, 2020c).</u> Papi et al. (2006) also indicate that chronic obstructive pulmonary disease (COPD) was significantly exacerbated by respiratory viral infections that cause reduction of forced expiratory volume in *1s* (FEV1) and airway inflammation (cf., Gorse et al., 2006). Ko et al. (2007) report that the most prevalent viruses detected during acute exacerbations of COPD in Hong Kong were the influenza A virus and coronavirus. They indicate that among patients with a mean age of more than 75 years, mean FEV1 was 40% of predicted normal and the FEV1/FVC (forced vital capacity) ratio was reduced to 58% of normal. De Serres et al. (2009) also suggested that the influenza virus frequently causes acute exacerbations of asthma and COPD. Moreover, the study by Wei et al. (2020) argues that air pollution in coastal city Weihai in China was slightly lower than the inland city of Jinan.<u>This study supports our results that the viral infectivity by COVID-19 is higher in hinterland cities rather than coastal cities in Italy.</u> Wei et al. (2020, p. 9) also suggest that different air quality strategies should be applied in inland and coastal cities: coastal cities need start bio aerosol risk alarm during moderate pollution when severe pollution occurs in inland cities.

Other studies have reported associations between air pollution and reduced lung function, increased hospital admissions, increased respiratory symptoms and high asthma medication use (Simoni et al., 2015; Jalaludin et al., 2004). In this context, the interaction between climate factors, air pollution and increased morbidity and mortality of people and children from respiratory diseases is a main health issue in society (Darrow et al., 2014). Asthma is a disease that has been associated with exposure to traffic-related air pollution and tobacco smoke (Liao, 2011). Many studies show that exposure to traffic-related outdoor air pollutants (e.g., particulate matter PM_10_ with an aerodynamic diameter ≤10 μm, nitrogen dioxide NO_2_, carbon monoxide CO, sulfur dioxide SO_2_, and ozone O_3_) increases the risk of asthma or asthma-like symptoms (Shankardass et al., 2009; Weinmayr et al., 2010). Especially, current evidence indicates that PM_10_ increases cough, lower respiratory symptoms and lower peak expiratory flow (Ward and Ayres, 2004; Nel, 2005). Weinmayr et al. (2010) provide strong evidence that PM_10_ may be an aggravating factor of asthma in children. Furthermore, asthma symptoms are exacerbated by air pollutants, such as diesel exhaust, PM_10_, NO_2_, SO_2_, and O_3_ and respiratory virus, such as adenovirus, influenza, parainfluenza and respiratory syncytial virus (Jaspers et al., 2005; Murdoch and Jennings, 2009; Murphy et al., 2000; Wong et al., 2009). The study by Liao et al. (2011) confirms that exacerbations of asthma have been associated with bacterial and viral respiratory tract infections and air pollution. Some studies have focused on the effect of meteorology and air pollution on acute viral respiratory infections and viral bronchiolitis (a disease linked to seasonal changes in respiratory viruses) in the first years of life (Nenna et al., 2017; Ségala et al., 2008; Vandini et al., 2013, 2015). Carugno et al. (2018) analyze respiratory syncytial virus (RSV), the primary cause of acute lower respiratory infections in children: bronchiolitis. Results suggest that seasonal weather conditions and concentration of air pollutants seem to influence RSV-related bronchiolitis epidemics in Italian urban areas. In fact, airborne particulate matter (PM) may influence the children’s immune system and foster the spread of RSV infection. This study also shows a correlation between short- and medium-term PM_10_ exposures and increased risk of hospitalization due to RSV bronchiolitis among infants. In short, manifold environmental factors–such as air pollution levels, circulation of respiratory viruses and colder temperatures–induce in longer periods of time spent indoors with higher opportunities for diffusion of infections between people. <u>In fact, in Italy the high diffusion of viral infectivity by COVID-19 in North of Italy is in winter period (February-March, 2020).</u> Studies also show that air pollution is higher during winter months and it has been associated with increased hospitalizations for respiratory diseases (Ko et al., 2007a; Medina-Ramón et al., 2006). Moreover, oscillations in temperature and humidity may lead to changes in the respiratory epithelium which increased susceptibility to infection (Deal et al., 1980). Murdoch and Jennings (2009) correlate the incidence rate of invasive pneumococcal disease (IPD) with fluctuations in respiratory virus activity and environmental factors in New Zealand, showing how incidence rates of IPD are associated with the increased activity of some respiratory viruses and air pollution. Another side effect of air pollution exposure is the association with the incidence of mumps. Hao et al. (2019) explore the effects of short-term exposure to air pollution on the incidence of mumps and show that exposure to NO_2_ and SO_2_ is significantly associated with higher risk of developing mumps. Instead, Yang et al. (2020) analyze the relationship between exposure to ambient air pollution and hand, foot, and mouth diseases (in short, HFMDs). Results show that the exposure of people to SO_2_, NO_2_, O_3_, PM_10_ and PM_2.5_ is associated with HFMDs. Moreover, the effect of air pollution in the cold season is higher than in the warm season. Shepherd and Mullins (2019) have also analyzed the relationship between arthritis diagnosis in those over 50 and exposure to extreme air pollution in utero or infancy. Results link early-life air pollution exposure to later-life arthritis diagnoses, and suggest a particularly strong link for Rheumatoid arthritis (RA)^3^. Sheperd and Mullins (2019) also argue that exposure to smog and air pollution in the first year of life is associated with a higher incidence of arthritis later in life. These findings are important to explain complex relationships between people, meteorological conditions, air pollution and viral infectivity because millions of people continue to be exposed to episodes of extreme air pollution each year in cities around the world.

### Air pollution, immune system and genetic damages

The composition of ambient particulate matter (PM) varies both geographically and seasonally because of the mix of sources at any location across time and space. A vast literature shows short-term effects of air pollution on health, but air pollution affects morbidity also in the long run (Brunekreef and Holgate, 2002). The mechanism of damages of air pollution on health can be explained as follows. Air pollutants exert their own specific individual toxic effects on the respiratory and cardiovascular systems of people; in addition, ozone, oxides of nitrogen, and suspended particulates have a common property of being potent oxidants, either through direct effects on lipids and proteins or indirectly through the activation intracellular oxidant pathways (Rahman and MacNee, 2000). Animal and human in-vitro and in-vivo exposure studies have demonstrated the powerful oxidant capacity of inhaled ozone with activation of stress signaling pathways in epithelial cells (Bayram et al., 2001) and resident alveolar inflammatory cells (Mochitate et al., 2001). Lewtas (2007) shows in human studies that exposures to combustion emissions and ambient fine particulate air pollution are associated with genetic damages. Long-term epidemiologic studies report an increased risk of all causes of mortality, cardiopulmonary mortality, and lung cancer mortality associated with increasing exposures to air pollution (cf., Coccia, 2012, 2014; Coccia and Wang, 2015). Although there is substantial evidence that polycyclic aromatic hydrocarbons or substituted polycyclic aromatic hydrocarbons may be causative agents in cancer and reproductive effects, an increasing number of studies–investigating cardiopulmonary and cardiovascular effects–shows potential causative agents from air pollution combustion sources.

About the respiratory activity, the adult lung inhales approximately 10-11,000 L of air per day, positioning the respiratory epithelium for exposure to high volumes of pathogenic and environmental insults. In fact, respiratory mucosa is adapted to facilitate gaseous exchange and respond to environmental insults efficiently, with minimal damage to host tissue. The respiratory mucosa consists of respiratory tract lining fluids; bronchial and alveolar epithelial cells; tissue resident immune cells such as alveolar macrophages (AM), dendritic cells, innate lymphoid cells and granulocytes; as well as adaptive memory T and B lymphocytes. In health, the immune system responds effectively to infections and neoplastic cells, with a response tailored to the insult, but must tolerate (i.e., not respond harmfully to) the healthy body and benign environmental influences. A well-functioning immune system is vital for a healthy body. Inadequate and excessive immune responses generate diverse pathologies, such as serious infections, metastatic malignancies and auto-immune conditions (Glencross et al., 2020). In particular, immune system consists of multiple types of immune cell that act together to generate (or fail to generate) immune responses. In this context, the explanation of relationships between ambient pollutants and immune system is vital to explain how pollution causes disease, and how pathology can be removed. Glencross et al. (2020) show that air pollutants can affect different immune cell types, such as particle-clearing macrophages, inflammatory neutrophils, dendritic cells that orchestrate adaptive immune responses and lymphocytes that enact those responses. In general, air pollutants stimulate pro-inflammatory immune responses across multiple classes of immune cell. Air pollution can enhance T helper lymphocyte type 2 and T helper lymphocyte type 17 adaptive immune responses, as seen in allergy and asthma, and dysregulate anti-viral immune responses. In particular, the association between high ambient pollution and exacerbations of asthma and chronic obstructive pulmonary disease (COPD) is consistent with immunological mechanisms. In fact, diseases can result from inadequate responses to infectious microbes allowing fulminant infections, inappropriate/excessive immune responses to microbes leading to more (collateral) damages than the microbe itself, and inappropriate immune responses to self/environment, such as seems to be in the case of COVID-19. Glencross et al. (2020) also discuss evidence that air pollution can cause disease by perturbing multicellular immune responses. Studies confirm associations between elevated ambient particulate matter and worsening of lung function in patients with COPD (Bloemsma et al., 2026), between COPD exacerbations and both ambient particulate matter and ambient pollutant gasses (Li et al., 2026) and similarly for asthma exacerbations with high concentration of ambient pollutants (Orellano et al., 2017, Zheng et al., 2015). In short, the associations between ambient pollution and airways exacerbations are stronger than associations with development of chronic airways diseases. Glencross et al. (2020) also argue that ambient pollutants can directly trigger cellular signaling pathways, and both cell culture studies and animal models have shown profound effects of air pollutants on every type of immune cell studied. In addition to the general pro-inflammatory nature of these effects, many of studies suggest an action of air pollution to augment Th2 immune responses and perturb antimicrobial immune responses. This mechanism also explains the association between high air pollution and increased exacerbations of asthma – a disease characterized by an underlying Th2 immuno-pathology in the airways with severe viral-induced exacerbations. Moreover, as inhaled air pollution deposits primarily on the respiratory mucosa, potential strategies to reduce such effects may be based on vitamin D supplementation. Studies show that plasma levels of vitamin D, activated by ultraviolet B, are significantly higher in summer and fall than winter and spring, in a latitude-dependent manner (Barger-Lux and Heaney, 2002). Since the temperature and hour of sun are dependent upon the latitude of population residence and influenced by urban/rural residence, Oh et al. (2010) argue that adequate activated vitamin D levels are also associated with diminished cancer risk and mortality (Lim et al., 2006; Grant, 2002). For instance, breast cancer incidence correlates inversely with the levels of serum vitamin D and ultraviolet B exposure, which are the highest intensity in summer season. These relationships of vitamin D and cancer risk are not limited to breast cancer, but are also relevant to colon, prostate, endometrial, ovarian, and lung cancers (Zhou et al., 2005).

In the context of this study and considering the negative effects of air pollution on human health and transmission dynamics of viruses, summer season may have twofold effects to reduce diffusion of viral infectivity:

1. hot and sunny weather increases temperature and improves environment that can reduce air pollution, typically of winter period, and as result alleviate transmission of viral infectivity by COVID-19 (Ko et al., 2007a; Medina-Ramón et al., 2006; Wei et al, 2020; Dowell et al., 2003; Kim et al., 1996; Talbot et al. 2005);
2. sunny days and summer season induce in population a higher production of vitamin D that reinforces and improves the function of immune system to cope with viral infectivity of COVID-19.

Overall, then, statistical analysis, supported by relevant studies in these research topics, reveals that accelerated transmissions dynamics of COVID-19 is also to air pollution-to-human transmission in addition to human-to-human transmission.

## PROACTIVE STRATEGIES TO PREVENT FUTURE EPIDEMICS SIMILAR TO COVID-19

At the end of 2019, medical professionals in Wuhan (China) were treating cases of pneumonia cases that had an unknown source (Li et al., 2020; Zhu and Xie, 2020; Chan et al., 2020; Backer et al., 2020). Days later, researchers confirmed the illnesses were caused by a new coronavirus (COVID-19). By January 23, 2020, Chinese authorities had shut down transportation going into and out of Wuhan, as well as local businesses, in order to reduce the spread of viral infectivity (Centers for Disease Control and Prevention, 2020; Public Health England. 2020; Manuell and Cukor, 2011). It was the first in the modern history of several quarantines set up in China and other countries around the world to cope with transmission dynamics of COVID-19. Quarantine is the separation and restriction of movement of people who have potentially been exposed to a contagious disease to ascertain if they become unwell, in order to reduce the risk of them infecting others (Brooks et al., 2019). In short, quarantine can generate a strong reduction of the transmission of viral infectivity. In the presence of COVID-19 outbreak in North Italy, Italian government has applied the quarantine and lockdown from 11 March, 2020 to 13 April, 2020 for all Italy, adding also some holidays thereafter. In fact, Italy was not able to prevent this complex problem of epidemics and has applied quarantine as a recovery strategy to lessen the health and socioeconomic damages caused by COVID-19. Millions of people have been quarantined for the first time in Italy and is one of the largest actions in the history of Italy. In addition, Italy applied non-pharmaceutical interventions based on physical distancing, school and store closures, workplace distancing, to avoid crowded places, similarly to the COVID-19 outbreak in Wuhan (cf., Prem et al., 2020). The benefits to support these measures until April, 2020 are aimed at delaying and reducing the height of epidemic peak, affording health-care systems more time to expand and respond to this emergency and, as a result reducing the final size of COVID-19 epidemic. In general, non-pharmaceutical interventions are important factors to reduce the epidemic peak and the acute pressure on the health-care system (Prem et al., 2020; Fong et al., 2020). However, Brooks et al. (2019) report: “negative psychological effects of quarantine including post-traumatic stress symptoms, confusion, and anger. Stressors included longer quarantine duration, infection fears, frustration, boredom, inadequate supplies, inadequate information, financial loss, and stigma. Some researchers have suggested long-lasting effects. In situations where quarantine is deemed necessary, officials should quarantine individuals for no longer than required, provide clear rationale for quarantine and information about protocols, and ensure sufficient supplies are provided. Appeals to altruism by reminding the public about the benefits of quarantine to wider society can be favourable”.

This strategy, of course, does not prevent future epidemics similar to the COVID-19 and it does not protect regions from future viral threats. Nations, alike Italy, have to apply *proactive strategies* that anticipate these potential problems and works to prevent them, reducing the health and economic impact in society.

### Suggested proactive strategies to prevent future epidemics similar to COVID-19

Daszak et al. (2020) argue that to prevent the next epidemic and pandemic similar to COVID-19, research and investment of nations should focus on:

surveillance among wildlife to identify the high-risk pathogens they carry
surveillance among people who have contact with wildlife to identify early spillover events
improvement of market biosecurity regarding the wildlife trade.

In addition, high surveillance and proper biosafety procedures in public and private institutes of virology that study viruses and new viruses to avoid that may be accidentally spread in surrounding environments with damages for population and vegetation. In this context, international collaboration among scientists is basic to address these risks, support decisions of policymakers to prevent future pandemic creating potential huge socioeconomic issues worldwide (cf., Coccia and Wang, 2016)^4^. In fact, following the COVID-19 outbreak, The Economist Intelligence Unit (EIU) points out that the global economy may contract of about by 2.2% and Italy by −7% of real GDP growth % in 2020 (EIU, 2020). Italy and other advanced countries should introduce organizational, product and process innovations to cope with future viral threats, such as the expansion of hospital capacity and testing capabilities, to reduce diagnostic and health system delays also using artificial intelligence, and as a consequence new ICT technologies for alleviating and/or eliminating effective interactions between infectious and susceptible individuals, and finally of course to develop effective vaccines and antivirals that can counteract future global public health threat in the presence of new epidemics similar to COVID-19 (Chen et al, 2020; Wilder-Smith et al 2020; Riou and Althaus, 2020; Yao et al., 2020; cf., Coccia, 2015, 2017, 2019, 2020).

This study here shows that geo-environmental factors of accelerated diffusion of COVID-19 are also likely associated with high air pollution and specific meteorological conditions (low wind speed, etc.) of North Italy and other Norther Italian regions that favor the transmission dynamics of viral infectivity. North Italy is one of the European regions with the highest motorization rate and polluting industrialization (cf., Legambiente, 2019). In 2018 in 55 provincial capitals the daily limits for PM_10_ or ozone were exceeded (i.e., 35 days for PM_10_ and 25 for ozone). In 24 of the 55 Italian province capitals, the limit was exceeded for both parameters, with negative effects on population that had to breathe polluted air for about four months in the year with subsequent health problems. In fact, the cities that last year passed the higher number of polluted days are Brescia with 150 days (47 for the PM_10_ and 103 for the ozone), followed by Lodi with 149 (78 for the PM_10_ and 71 for the ozone),–these are two cities with severe COVID-19 outbreak–, Monza (140), Venice (139), Alessandria (136), Milan (135), Turin (134), Padua (130), Bergamo and Cremona (127) and Rovigo (121). These provincial capitals of the River Po area in Italy have exceeded at least one of the two limits just mentioned. The first city not located in the Po valley is Frosinone (Lazio region of the central part of Italy) with 116 days of exceedance (83 for the PM_10_ and 33 for the ozone), followed by Genoa with 103 days, Avellino a city close to Naples in South Italy (Campania region with 89 days: 46 for PM_10_ and 43 for ozone) and Terni with 86 (respectively 49 and 37 days for the two pollutants). Many cities in Italy are affected by air pollution and smog because of traffic, domestic heating, industries and agricultural practices and with private cars that continue to be by far the most used means of transportation (more than 39 million cars in 2019). In fact, a major source of emissions of nitrogen oxides into the atmosphere is the combustion of fossil fuels from stationary sources (heating, power generation) and motor vehicles. In ambient conditions, nitric oxide is rapidly transformed into nitrogen dioxide by atmospheric oxidants such as ozone (cf., Brunekreef and Holgate, 2002). In Italy, the first COVID-19 outbreak has been found in Codogno, a small city of the Lodi area, close to Milan. Although local lockdown as red zone on February 25, 2020, the Regional Agency for Environmental Protection showed that concentrations of PM_10_ beyond the limits in almost all of Lombardy region including the red zone (i.e., 82 mm/m^3^ of air measured in Codogno). The day after, February 26, 2020, the mistral wind and then the north wind swept the entire Po valley, bringing to Lombardy region a substantial reduction in the average daily concentrations of PM_10_, which almost everywhere were lower than 50 micrograms of particulate matter/m^3^ of air.

Hence, high concentration of nitrogen dioxide, a noxious gas, particulate air pollutants emitted by motor vehicles, power plants, and industrial facilities in North Italy seems to be a platform to support diffusion of viral infectivity (Groulx et al., 2018), increase hospitalizations for respiratory virus bronchiolitis (cf., Carugno et al., 2018; Nenna et al., 2017), increase asthma incidence (Liao et al., 2011) and damage to the immune system of people (Glencross et al., 2020). Transmission dynamics of COVID-19 has found in air pollution and meteorological conditions of North Italy an appropriate environment and population to carry out an accelerated diffusion that is generation more than 13,000 deaths and a huge number of hospitalizations in a short period of time.

An *indirect effect* of quarantine and lockdown in Italy is the strong reduction of airborne Nitrogen Dioxide Plummets and PM_10_ over Norther of Italy. The maps in figure 4 by ESA (2020) show concentrations of nitrogen dioxide NO_2_ values across Italy before the quarantine and lockdown in February, 2020 and during the quarantine and lockdown in March, 2020. The reduction in NO_2_ pollution is apparent in all North Italy (Po Valley). Hence, the measures taken to cope with the COVID-19 outbreak (closure of schools and the reduction of traffic), particularly restrictive in the first phase on the regions of Northern Italy, have allowed a drastic reduction of concentrations of fine particulate matter, nitrogen dioxide and other polluting substances on the Po Valley. For instance, in Piedmont, one of the regions of North Italy also having a high COVID-19 diffusion, the concentration of air pollution since the beginning of March, 2020 has ever exceeded the limit values of PM_10_ and has always remained below 50µg / m^3^ everywhere. Overall, then, the indirect effect of quarantine and lockdown of Italy and other European countries has reduced in a short time NO_2_ and air pollution, improving the quality of environment that may reduce, associated with quarantine, physical distancing and other inter-related factors, the transmission dynamics of COVID-19. A study by Zhang et al. (2019a) shows that with the implementation of air policy in China, from 2013 to 2017, fine particle (PM_2.5_) concentrations have significant declined nationwide with health benefits. Now, the danger is that after the quarantine and lockdown, the industrial activity of industrialized regions in Italy has to resume at an intense pace of production and in next winter-fall season 2020-2021 there may be again the environmental and meteorological conditions that can lead to diffusion of viral infectivity of COVID-19 and/or other dangerous viruses. <u>Of course, non-physical distancing and other long-run factors play a critical part in mitigating transmission dynamics of future epidemic similar to COVID-19, in particular when measures of physical distancing, school and store closures, workplace distancing, prohibition for crowded places are relaxed.</u> The suggested strategy that regions of North Italy has to apply, considering their geographical locations and meteorological conditions with a high density of polluting industrialization, is to avoid to overcome the limits set of PM_10_ and other pollutants, following more and more a sustainable pathways of growth. *One of the findings here suggests that the max number of days per year that Italian provincial capitals can exceed the limits set for PM*_*10*_ (*particulate matter 10 micrometers or less in diameter*) *or for ozone, considering the meteorological condition has to be less than 50 days. After this critical point, the study suggests that environmental inconsistencies because of the combination between air pollution and meteorological conditions trigger a take-off of viral infectivity* (*epidemic diffusion*) *with damages for health of population and economy in society*. Italy must design and set up necessary measures to drastically reduce the concentrations of pollution present and improve air quality in cities. Italy not has to respect Legislative Decree 155/2010 that establishes a maximum number of 35 days / year with concentrations higher than 50 μg / m^3^. As a matter of fact, the quarantine and other non-pharmaceutical interventions can reduce the impact of viral infectivity in the short term, but to prevent future epidemics similar to COVID-19, Italy and advanced nations have more and more to sustain a sustainable growth. The environmental policy has to be associated with sustainable technologies that reduce air pollution improving the quality of air and environment for population to cope with future viral threats (cf., Coccia, 2005, 2006, 2018; Coccia and Watts, 2020)^5^. Italy must support, more and more, sustainable mobility as engine of socioeconomic change and redesign cities for people using an urban planning that improves public respiratory health. Moreover, in the presence of the association between air pollution, climate^6^ and viral infectivity. Italy and other advanced nations have to immediately reduce the motorization rate of polluting machines with a transition to new electric vehicles, generating a revolution in society. It is basic to encourage sustainable mobility, by enhancing local, urban and commuter public transport with electric vehicles and creating vast Low Emission Zones within cities. Italy has to launch a real sustainable growth roadmap with the aim of complete zero emissions in all socioeconomic system. Some studies done in the past show the causality of the reduction of air pollution on health benefits. For instance, Pope (1989) describes the case of a labor dispute that shut down a large steel mill in the Utah Valley for 14 months in 1987. Toxicological studies of particulate matter collected before, during, and after the strike, in the Utah Valley case, provide strong evidence of a causal relation between exposure to ambient particulate matter and mortality and morbidity. Ambient particulate matter concentrations as well as respiratory hospital admissions were clearly decreased during the strike, increasing to prestrike levels after the dispute ended (Pope, 1989; cf., the reduction of mortality described by Pope, 1996). Another example includes the reductions in acute-care visits and hospital admissions for asthma in Atlanta (GA, USA), in conjunction with reduced air pollution due to traffic restrictions taken during the 2000 Olympic games (Friedman, 2001).

**Figure 4.**
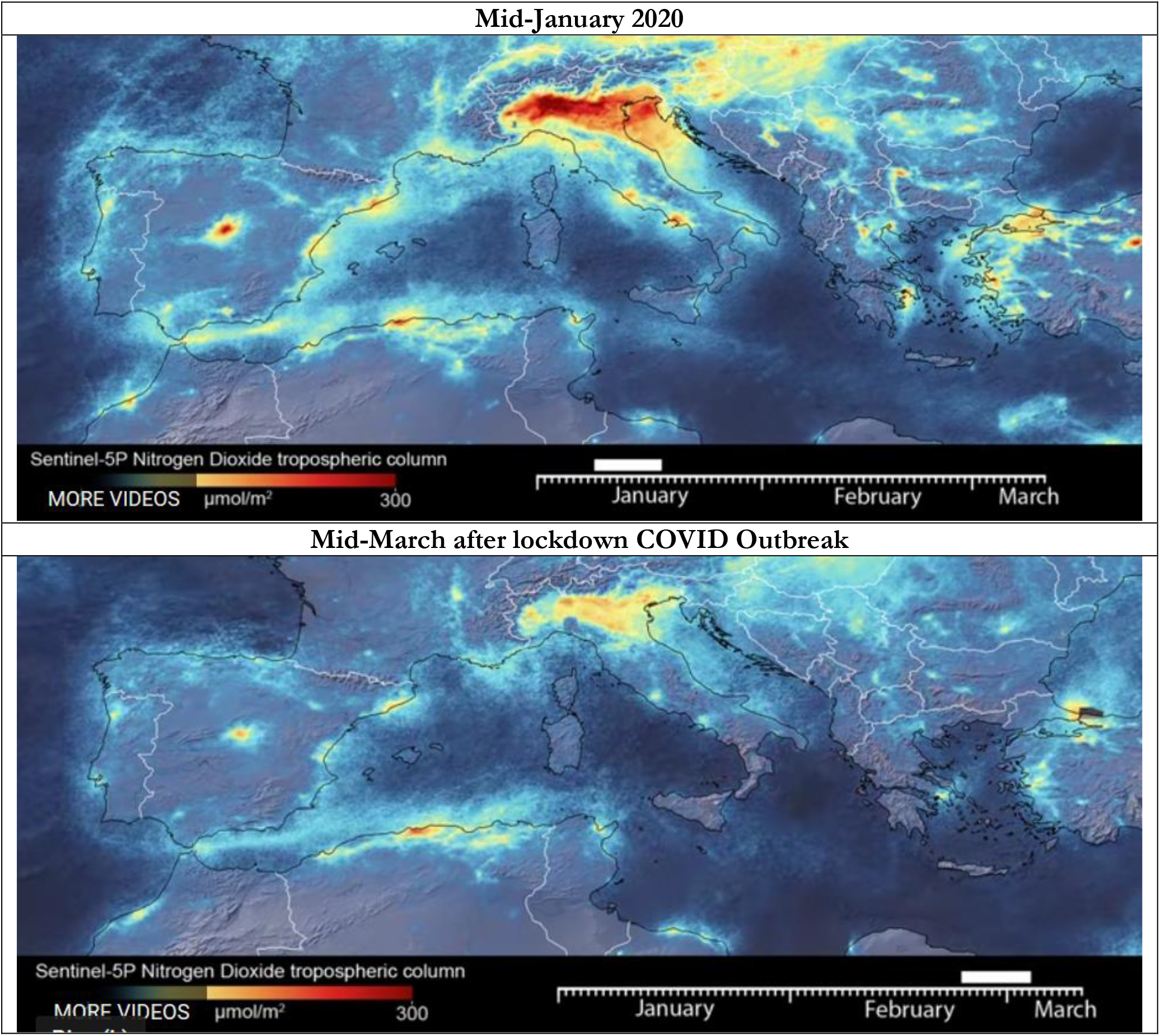
Reduction Nitrogen Dioxide before and after lockdown COVID Outbreak in Italy. *Sources*: European Space Agency, 2020.

## CONCLUSIONS

The intensity of human interactions with Earth systems has accelerated in recent decades, because of urban development, population growth, industrialization, deforestation, construction of dams, etc., with changes in physical, biological, and chemical processes in soils and waters. In particular, human activity, driven by a high level of world population that is about eight billion (U. S. Census Bureau 2020), has induced changes to Earth’s surface, cryosphere, ecosystems, and climate that are now so great and rapid, advancing the geological epoch of Anthropocene (Crutzen and Stoermer, 2000; Foley et al., 2013). The beginning of the Anthropocene at around 1780 AD marks the beginning of immense rises in human population and carbon emissions as well as atmospheric CO^2^ levels (Ellis et al., 2013). The scale of carbon emissions associated with industrial activity is leading to a rise in atmospheric greenhouse gases at a rate unprecedented and gradual rise in carbon dioxide (Glikson, 2013; Coccia, 2014a). In this era of Anthropocene, the health effects of air pollution have been subject to intense study in recent years. Exposure to airborne particulate matter and ozone has main health effects associated with increases in mortality and hospital admissions for respiratory and cardiovascular diseases (Kampa and Castanas, 2008; Hoek et al., 2013). The idea that air pollution episodes have a detrimental effect on health is now rarely contested, and acute exposures to high concentrations of air pollutants exacerbate cardiopulmonary disorders in human population worldwide (Langrish and Mills, 2014).

This study shows that factors determining the diffusion of epidemics similar to COVID-19 are due to manifold elements, in addition to human-to-human transmission, given by:

1. *General factors* that are the same for all locations and associated with innate biological characteristics of the viruses, incubation time, effects on infected and susceptible people, etc.
2. *Specific factors* that are different for each location and even for each individual, such us level of air pollution over time and space, meteorological conditions of specific location, season, density of areas, economic wealth, cultural characteristics (religious habits, food culture, etc.), organization and efficiency of healthcare sector, facilities and equipment in health sector, immune system of people, average age of population, sex of people, etc.

The main results of the study here, based on case study of COVID-19 outbreak in Italy, are:

○ The acceleration of transmission dynamics of COVID-19 in North Italy has a high association with air pollution of cities measured with days of exceeding the limits set for PM10 or ozone
○ Cities having more than 100 days of air pollution (exceeding the limits set for PM_10_), they have a very high average number of infected individual (about 3,100 infected), whereas cities having less than 100 days of air pollution, they have a lower average number of infected (about 900 infected individuals)
○ Hinterland cities with higher number of average days exceeding the limits set for PM_10_ have a very high number of infected people on 1^st^ April, 2020 (arithmetic mean is about 2,000 infected, with average polluted days more than 80), than coastal cities also having days of exceeding the limits set for PM10 or ozone (arithmetic mean about 700 infected, with average polluted days about 60). In fact, coastal cities have an average higher intensity of wind speed (about 12 km/h) than hinterland cities (8 km/h) and statistical analysis reveals a negative coefficient correlation between number of infected and intensity of wind speed (*r*= –28 to –38%, *p*-value <0.05): in fact, wind speed and other elements clean air from pollutants that are associated with transmission dynamics of viral infectivity.
○ Air pollution in cities under study seems to be a more important predictor in the initial phase of transmission dynamics (on 17^th^ March 2020, b_1_ = 1.27, *p*<0.001) than human-to-human transmission (b_2_ = 0.31, *p*<0.05). In the second phase of the transmission dynamics of viral infectivity, air pollution reduces intensity (on 1^st^ April, 2020 b’_1_ = .85, *p*<0.001) also because of indirect effect of lockdown and human-to-human transmission slightly increases (b’_2_ = 0.34, *p*<0.01): *This result reveals that accelerated transmissions dynamics of COVID-19 is due to mainly air pollution-to-human transmission in addition to human-to-human transmission*.
○ To minimize future epidemics similar to COVID-19, the max number of days per year in which Italian provincial capitals can exceed the limits set for PM_10_ (particulate matter 10 micrometers or less in diameter) or for ozone, considering their meteorological conditions, is about 45 days.

Hence, high concentration of nitrogen dioxide, a noxious gas, particulate air pollutants emitted by motor vehicles, power plants, and industrial facilities in North Italy seems to be a platform to support diffusion of viral infectivity (Groulx et al., 2018), increase hospitalizations for respiratory virus bronchiolitis (cf., Carugno et al., 2018; Nenna et al., 2017), increase asthma incidence (Liao et al., 2011) and damage to the immune system of people (Glencross et al., 2020). Beelen et al. (2013) report the need to draw attention to the continuing effects of air pollution on health. A socioeconomic strategy to prevent future epidemics similar to the COVID-19 is also the reduction of pollution with fruitful environmental and health effect by the rationalization of manufacturing industry in a perspective of sustainable development, de-industrializing polluting activities in the geographical development of current capitalism. De-industrialization of polluting industries and sustainable development impose often huge social costs in the short term on people, households, and families but they have long-run benefits for human societies. Studies show that public and environmental health policy interventions are necessary and have the potential to reduce morbidity and mortality across Europe (cf., Raaschou-Nielsen et al., 2013). In fact, the improvements in air quality have been accompanied by demonstrable benefits to human health. Pope et al. (2009) reported that PM_2.5_ concentrations fell by a third from the early 1980s to the late 1990s across major US metropolitan areas, with each 10 μg/m^3^ reduction associated with an increase in life expectancy of 0.61 years. Because of health problems of polluting industrialization, Wei et al. (2020) suggest different air pollution regulations in regions having varied geographical and climatic conditions, and different bio aerosol pollution. In particular, Wei et al. (2020) suggest that different air quality strategies should be applied in inland and coastal cities, e.g., coastal cities also need start bio aerosol risk alarm during moderate pollution when severe pollution occurs in inland cities. Guo et al. (2019) argue that in recent years, haze pollution is a serious environmental problem affecting cities, proposing implications for urban planning to improve public respiratory health. In short, the long-term benefits of sustainable economic development are basic for the improvement of environment, atmosphere, air quality and especially health of populations (Blackaby, 1978; Bluestone and Harrison, 1982; Pike, 2009).

Overall, the, these findings here are consistent with correlational studies and indicate that health effects of air pollution exposure can span decades and extend beyond cardiopulmonary systems affecting diffusion of epidemics similar to COVID-19. Hence, it is important to reinforce evidence related to air pollution and inter-related factors of the transmission dynamics of virus similar to COVID-19, and helps policy makers to develop proactive regulations for the control of environment, air pollution, polluting industrialization and prevention of the diffusion of viral infectivity. The complex problem of epidemic threats has to be treated with an approach of dissolution: it means to redesign the strategies and protocols to cope with future epidemics in such way as to eliminate the conditions that caused accelerated diffusion of COVID-19, thus enabling advanced nations to do better in the future than the best it can do today (Ackoff and Rovin, 2003, pp. 9-10; Bundy et al., 2017). This study revels interesting results of transmission dynamics of COVID-19 given by the mechanism of air pollution-to-human transmission that in addition to human-to-human transmission seems to have accelerated diffusion of epidemics in Italy. However, these conclusions are tentative. There are several challenges to such studies, particularly in real time. Sources may be incomplete, or only capture certain aspects of the on-going outbreak dynamics; there is need for much more research in to the relations between viral infectivity, air pollution, meteorological factors and other determinants, when the COVID-19 outbreak is over. Overall, then, in the presence of polluting industrialization of cities and air pollution - to-human transmission of viral infectivity, this study must conclude that a comprehensive strategy to prevent future epidemics similar to COVID-19 has also to be designed in environmental and socioeconomic terms, that is in terms of sustainability science and environmental science, and not only in terms of biology, healthcare and health sector.

## Data Availability

Data are available to the following link:
https://lab24.ilsole24ore.com/coronavirus/

## Footnotes

1 Flint (1884, p. 105, original emphasis): “Vico entirely accepted. *Vere scire est per causas scire* like Aristotle and every one else who has undertaken to defend the position that there can be no adequate knowledge where causes are unknown, he had to understand the word cause in a wide sense, inclusive of conditions and principles in general as well as of causes strictly so termed. The cause of a thing is whatever adequately explains it,—the whole ground, reason, or source of it, *quella che per produrre l’effetto non ha d’altra bisogno* Hence also the phrases, ‘to prove by causes,’ ‘to collect the elements of a thing,’ and ‘to make,’ are understood as equivalent: *probare per causas idem est ac efficere, —probare a causis est elementa rei colligere*.”

2 High-efficiency particulate air, also known as high-efficiency particulate absorbing and high-efficiency particulate arrestance, is an efficiency standard of air filter.

3 Rheumatoid arthritis is a chronic inflammatory disorder in which the body’s immune system attacks its joints, and is one of the most common autoimmune diseases (Cooper and Stroehla, 2003). Moreover, rheumatoid arthritis is a major cause of disability that reduces patient’s lifespan by 15-20% from the onset of the illness (Myllykangas-Luosujäarvi et al., 1995; cf., Chang et al., 2016; De Roos et al., 2014; Farhat et al., 2011; Jung et al., 2017).

4 Socioeconomic shocks can lead to a general increase of prices, high public debts, high unemployment, income inequality and as a consequence violent behavior (Coccia, 2016, 2017, 2017a).

5 cf. for dynamics of technological and economic change in society the studies by Coccia (2005a, 2005b, 2008, 2008, 2009, 2015a, 2017e, 2017f, 2017g, 2018a, 2019a, 2019b, 2019c, 2019d; Coccia and Finardi, 2012; Coccia and Rolfo, 2008).

6 Some studies show that in addition to human-to-human contact, ambient temperature is an important factor in the transmission and survival of coronaviruses (Zhu et al., 2020) as well as temperature variation and humidity may also be important factors affecting the COVID-19 mortality (Ma et al., 2020)

## Notes

### Competing Interest Statement

The authors have declared no competing interest.

## REFERENCES

Ackoff R. L., Rovin S. 2003. Redesigning Society, Stanford University Press, Stanford, CA.

Backer J.A., Klinkenberg D., Wallinga J. 2020. Incubation period of 2019 novel coronavirus (2019-nCoV) infections among travellers from Wuhan, China, 20–28 January 2020. Euro Surveill 2020;25: 2000062.

Barger-Lux MJ, Heaney RP 2002. Effects of above average summer sun exposure on serum 25-hydroxyvitamin D and calcium absorption. J Clin Endocrinol Metab 87(11):4952–4956

Bayram H., Sapsford R.J., Abdelaziz M.M., Khair O.A. 2001. Effect of ozone and nitrogen dioxide on the release of proinflammatory mediators from bronchial epithelial cells of nonatopic nonasthmatic subjects and atopic asthmatic patients in vitro. J Allergy Clin Immunol; 107: 287–94.

Beelen R., Raaschou-Nielsen O., Stafoggia M., et al. Effects of long-term exposure to air pollution on natural-cause mortality: an analysis of 22 European cohorts within the multicentre ESCAPE project. Lancet 2013; Published online Dec 9. http://dx.doi.org/10.1016/S0140-6736(13)62158-3.

Bell M.L., Davis D.L., Fletcher T., 2004. A retrospective assessment of mortality from the London Smog episode of 1952: the role of influenza and pollution. Environ. Health Perspect. 112 (1), 6.

Blackaby F. 1978. De-Industrialisation. London: Heinemann.

Bloemsma L.D., G. Hoek, L.A. Smit, Panel studies of air pollution in patients with COPD: systematic review and meta-analysis, Environ. Res. 151 (2016) 458–468.

Bluestone B., Harrison B. 1982. The Deindustrialization of America: Plant Closings, Community Abandonment and the Dismantling of Basic Industry. New York: Basic Books.

Brooks S. K, Webster R. K., Smith L. E., Woodland L., Wessely S., Greenberg N., Rubin G. J. 2019. The psychological impact of quarantine and how to reduce it: rapid review of the evidence. The lance5t, rapid review, Published Online February 26, 2020, https://doi.org/10.1016/S0140-6736(20)30460-8

Brunekreef B., Holgate S. T. 2002. Air pollution and health, Lancet 2002; 360: 1233–42

Brunekreef B., Holgate S. T. 2002. Air pollution and health, The Lancet, vol. 360, n. 9341, pp. 1233-1242, https://doi.org/10.1016/S0140-6736(02)11274-8.

Bundy J., Pfarrer M. D., Short C. E., Coombs W. T. 2017. Crises and Crisis Management: Integration, Interpretation, and Research Development. Journal of Management, 43 (6): 1661–1692. doi:10.1177/0149206316680030

Camacho A., Kucharski A., Aki-Sawyerr Y. et al. 2015. Temporal changes in Ebola transmission in Sierra Leone and implications for control requirements: a real-time modelling study. PLoS Curr 2015; 7.

Carugno M., Dentali F., Mathieu G., Fontanella A., Mariani J., Bordini L., Milani G. P., Consonni D., Bonzini M., Bollati V., Pesatori A. C. 2018. PM10 exposure is associated with increased hospitalizations for respiratory syncytial virus bronchiolitis among infants in Lombardy, Italy, Environmental Research 166 (2018) 452–457, https://doi.org/10.1016/j.envres.2018.06.016

Centers for Disease Control and Prevention, 2020. Quarantine and isolation. 2017. https://www.cdc.gov/quarantine/index.html (Accessed Jan 30, 2020).

Chan J.F.W., Yuan S., Kok K.H., et al. 2020. A familial cluster of pneumonia associated with the 2019 novel coronavirus indicating person-to-person transmission: a study of a family cluster. Lancet 2020; 395: 514–23.

Chang K.H., Hsu, C.C., Muo C.H., Hsu C.Y., Liu H.C., Kao C.H., Chen C.Y., Chang M.Y., Hsu Y.C. 2016. Air pollution exposure increases the risk of rheumatoid arthritis: a longitudinal and nationwide study. Environ. Int. 94, 495–499.

Charmi H., Sneha G., Ujwalkumar T., 2018. A review on recent progress in observations, and health effects of bio aerosols. Environ. Int. 118, 189e193.

Chen S., Yang J., Yang W., Wang C, Barnighausen T. 2020. COVID-19 control in China during mass population movements at New Year. Lancet; 395: 764–66.

Churchman C. W. 1971. The design of inquiring systems. Basic Books, New York.

Coccia M. 2005. Metrics to measure the technology transfer absorption: analysis of the relationship between institutes and adopters in northern Italy. International Journal of Technology Transfer and Commercialization, vol. 4, n. 4, pp. 462–486. https://doi.org/10.1504/IJTTC.2005.006699

Coccia M. 2005a. A taxonomy of public research bodies: a systemic approach, Prometheus –The journal of issues in technological change. Innovation. Information economics, communications and science policy, vol. 23, n. 1, pp. 63–82. DOI:10.1080/0810902042000331322

Coccia M. 2005b. Countrymetrics: valutazione della performance economica e tecnologica dei paesi e posizionamento dell’Italia,

Rivista Internazionale di Scienze Sociali, vol. CXIII, n. 3, pp. 377-412. Stable URL: http://www.jstor.org/stable/41624216.

Coccia M. 2006. Analysis and classification of public research institutes, World Review of Science, Technology and Sustainable Development, vol. 3, n. 1, pp.1–16. https://doi.org/10.1504/WRSTSD.2006.008759

Coccia M. 2008. Spatial mobility of knowledge transfer and absorptive capacity: analysis and measurement of the impact within the geoeconomic space, The Journal of Technology Transfer, vol. 33, n. 1, pp. 105–122. https://doi.org/10.1007/s10961-007-9032-4

Coccia M. 2009. Research performance and bureaucracy within public research labs, Scientometrics, vol. 79, n. 1, pp. 93–107. https://doi.org/10.1007/s11192-009-0406-2

Coccia M. 2012. Driving forces of technological change in medicine: Radical innovations induced by side effects and their impact on society and healthcare, Technology in Society, vol. 34, n. 4, pp. 271–283, https://doi.org/10.1016/j.techsoc.2012.06.002

Coccia M. 2014. Path-breaking target therapies for lung cancer and a far-sighted health policy to support clinical and cost effectiveness, Health Policy and Technology, xvol. 1, n. 3, pp. 74–82. https://doi.org/10.1016/j.hlpt.2013.09.007

Coccia M. 2014a. Steel market and global trends of leading geo-economic players. International Journal of trade and global markets, vol. 7, n.1, pp. 36–52, DOI: http://dx.doi.org/10.1504/IJTGM.2014.058714

Coccia M. 2015. General sources of general purpose technologies in complex societies: Theory of global leadership-driven innovation, warfare and human development, Technology in Society, vol. 42, August, pp. 199–226, http://doi.org/10.1016/j.techsoc.2015.05.008

Coccia M. 2015a. Technological paradigms and trajectories as determinants of the R&D corporate change in drug discovery industry. Int. J. Knowledge and Learning, vol. 10, n. 1, pp. 29–43. http://dx.doi.org/10.1504/IJKL.2015.071052

Coccia M. 2016. The relation between price setting in markets and asymmetries of systems of measurement of goods, The Journal of Economic Asymmetries, vol. 14, part B, November, pp. 168–178, https://doi.org/10.1016/j.jeca.2016.06.001

Coccia M. 2017. Asymmetric paths of public debts and of general government deficits across countries within and outside the European monetary unification and economic policy of debt dissolution, The Journal of Economic Asymmetries, vol. 15, June, pp. 17–31, https://doi.org/10.1016/j.techfore.2010.02.003

Coccia M. 2017a. A Theory of general causes of violent crime: Homicides. Income inequality and deficiencies of the heat hypothesis and of the model of CLASH, Aggression and Violent Behavior, vol. 37, November-December, pp. 190–200, https://doi.org/10.1016/j.avb.2017.10.0051552, https://doi.org/10.1016/j.jengtecman.2019.11.003

Coccia M. 2017b. Sources of technological innovation: Radical and incremental innovation problem-driven to support competitive advantage of firms. Technology Analysis & Strategic Management, vol. 29, n. 9, pp. 1048–1061, https://doi.org/10.1080/09537325.2016.1268682

Coccia M. 2017c. Varieties of capitalism’s theory of innovation and a conceptual integration with leadership-oriented executives: the relation between typologies of executive, technological and socioeconomic performances. Int. J. Public Sector Performance Management, Vol. 3, No. 2, pp. 148–168. https://doi.org/10.1504/IJPSPM.2017.084672

Coccia M. 2017d. Asymmetric paths of public debts and of general government deficits across countries within and outside the European monetary unification and economic policy of debt dissolution, The Journal of Economic Asymmetries, vol. 15, June, pp. 17–31, https://doi.org/10.1016/j.techfore.2010.02.003

Coccia M. 2017e. Sources of disruptive technologies for industrial change. L’industria –rivista di economia e politica industriale, vol. 38, n. 1, pp. 97–120, DOI: 10.1430/87140

Coccia M. 2017f. The Fishbone diagram to identify, systematize and analyze the sources of general purpose technologies. Journal of Social and Administrative Sciences, http://dx.doi.org/10.1453/jsas.v4i4.1518

Coccia M. 2017g. The source and nature of general purpose technologies for supporting next K-waves: Global leadership and the case study of the U.S. Navy’s Mobile User Objective System, Technological Forecasting & Social Change, vol. 116 (March), pp. 331–339. https://doi.org/10.1016/j.techfore.2016.05.019

Coccia M. 2018. Theorem of not independence of any technological innovation, Journal of Economics Bibliography, vol. 5, n. 1, pp. 29–35, http://dx.doi.org/10.1453/jeb.v5i1.1578

Coccia M. 2018a. The origins of the economics of Innovation, Journal of Economic and Social Thought, vol. 5, n. 1, pp. 9–28, http://dx.doi.org/10.1453/jest.v5i1.1574

Coccia M. 2019. Why do nations produce science advances and new technology? Technology in society, vol. 59, November, 101124, pp. 1–9, https://doi.org/10.1016/j.techsoc.2019.03.007

Coccia M. 2019a. Comparative Institutional Changes. A. Farazmand (ed.), Global Encyclopedia of Public Administration, Public Policy, and Governance, Springer Nature Switzerland AG, https://doi.org/10.1007/978-3-319-31816-5_1277-1

Coccia M. 2019b. Comparative Theories of the Evolution of Technology. In: Farazmand A. (eds) Global Encyclopedia of Public Administration, Public Policy, and Governance. Springer, Cham, Springer Nature Switzerland AG, https://doi.org/10.1007/978-3-319-31816-5_3841-1

Coccia M. 2019c. Comparative World-Systems Theories. A. Farazmand (ed.), Global Encyclopedia of Public Administration, Public Policy, and Governance, Springer Nature Switzerland AG, https://doi.org/10.1007/978-3-319-31816-5_3705-1

Coccia M. 2019d. The Role of Superpowers in Conflict Development and Resolutions. A. Farazmand (ed.), Global Encyclopedia of Public Administration, Public Policy, and Governance, Springer Nature Switzerland AG, https://doi.org/10.1007/978-3-319-31816-5_3709-1

Coccia M. 2020. Deep learning technology for improving cancer care in society: New directions in cancer imaging driven by artificial intelligence. Technology in Society, vol. 60, February, pp. 1–11, https://doi.org/10.1016/j.techsoc.2019.101198.

Coccia M., Finardi U. 2012. Emerging nanotechnological research for future pathway of biomedicine. International Journal of Biomedical nanoscience and nanotechnology, vol. 2, nos. 3-4, pp. 299–317. DOI: 10.1504/IJBNN.2012.051223

Coccia M., Rolfo S. 2008. Strategic change of public research units in their scientific activity, Technovation, vol. 28, n. 8, pp. 485–494. https://doi.org/10.1016/j.technovation.2008.02.005

Coccia M., Wang L. 2015. Path-breaking directions of nanotechnology-based chemotherapy and molecular cancer therapy, Technological Forecasting & Social Change, 94(May):155–169. https://doi.org/10.1016/j.techfore.2014.09.007

Coccia M., Wang L. 2016. Evolution and convergence of the patterns of international scientific collaboration, Proceedings of the National Academy of Sciences of the United States of America, vol. 113, n. 8, pp. 2057–2061, www.pnas.org/cgi/doi/10.1073/pnas.1510820113.

Coccia M., Watts J. 2020. A theory of the evolution of technology: technological parasitism and the implications for innovation management, Journal of Engineering and Technology Management, vol. 55 (2020) 101552, S0923-4748(18)30421-1, https://doi.org/10.1016/j.jengtecman.2019.11.003

Cooper BS, Pitman RJ, Edmunds WJ, Gay NJ. 2006. Delaying the international spread of pandemic influenza. PLoS Med 2006; 3: e212.

Cooper, G.S., Stroehla, B.C., 2003. The epidemiology of autoimmune diseases. Autoimmun. Rev. 2 (3), 119–125.

Crutzen P.J., Stoermer E.F., 2000. The ‘‘Anthropocene’’. IGBP Newsletter 41 (1) 17–18.

Darrow, L.A., Mitchel, K., Flanders, W.D., Mulholland, J.A., Tolbert, P.E., Strickland, M.J., 2014. Air pollution and acute respiratory infections among children 0–4 years of age: an 18-year time-series study. Am. J. Epidemiol. 180, 968–977. http://dx.doi.org/10.1093/aje/kwu234.

Das, P., Horton, R., 2017. Pollution, health, and the planet: time for decisive action. Lancet 391, 407–408.

Daszak P., Olival K. J., Li H. 2020. A strategy to prevent future epidemics similar to the 2019-nCoV outbreak, Biosafety and Health, http://dx.doi.org/10.1016/j.bsheal.2020.01.003

De Roos A.J., Koehoorn M., Tamburic L., Davies H.W., Brauer M. 2014. Proximity to traffic, ambient air pollution, and community noise in relation to incident rheumatoid arthritis. Environ. Health Perspect. 122 (10), 1075.

De Serres G., Lampron N., La Forge J., Rouleau I., Bourbeau J., Weiss K. et al. 2009. Importance of viral and bacterial infections in chronic obstructive pulmonary disease exacerbations. J Clin Virol; 46:129–33.

Deal E.C., McFadden E.R., Ingram R.H., Breslin F.J., Jaeger J.J. 1980. Airway responsiveness to cold air and hyperpnea in normal subjects and in those with hay fever and asthma. Am J Respir Dis; 121:621–8.

Després V., Huffman J.A., Burrows S.M., Hoose C., Safatov A., Buryak G., et al., 2012. Primary biological aerosol particles in the atmosphere: a review. Tellus B 64, 145–153.

Dong E., Du H., Gardner L. 2020. An interactive web-based dashboard to track COVID-19 in real time https://doi.org/10.1016/S1473-3099(20)30120-1

Dowell S.F., Whitney C.G., Wright C., Rose C.E., Schuchat A. 2003. Seasonal patterns of invasive pneumococcal disease. Emerging Infect Dis 2003; 9:573e9.

EIU 2020. COVID-19 to send almost all G20 countries into a recession, 26th Mar 2020.

Ellis E.C., Kaplan J.O., Fuller D.Q., Vavrus S., Goldewijk K.K., Verburg P.H. 2013. Used planet: a global history. Proceedings of the National Academy of Sciences, http://dx.doi.org/10.1073/pnas.1217241110

ESA (2020). European Space Agency Coronavirus: nitrogen dioxide emissions drop over Italy, www.esa.int, Mar 13, 2020 https://www.esa.int/ESA_Multimedia/Videos/2020/03/Coronavirus_nitrogen_dioxide_emissions_drop_over_Italy (Accessed march 2020)

European Centre for Disease Prevention and Control. 2020. Public health management of persons having had contact with novel coronavirus cases in the European Union. European Centre for Disease Prevention and Control, 2020. https://www.ecdc.europa.eu/en/publications-data/public-health-management-personshaving-had-contact-novel-coronavirus-cases (Accessed Feb 2, 2020).

European Centre for Disease Prevention and Control. 2020a. Risk assessment guidelines for diseases transmitted on aircraft. Part 2: Operational guidelines for assisting in the evaluation of risk for transmission by disease. 2011. https://www.ecdc.europa.eu/sites/default/files/media/en/publications/Publications/1012_GUI_RAGIDA_2.pdf (Accessed Feb 6, 2020).

Farhat S.C., Silva C.A., Orione M.A., Campos L.M., Sallum A.M., Braga A.L., 2011. Air pollution in autoimmune rheumatic diseases: a review. Autoimmun. Rev. 11 (1), 14–21.

Flint R. 1884. Vico, William Blackwood and sons, Edinburgh and London.

Foley S. F., Gronenborn D., Andreae M. O., Kadereit J.W., Esper J., Scholz D., Pöschl U., Jacob D. E., Schöne B. R., Schreg R., Vött A., Jordan D., Lelieveld J., Weller C. G., Alt K. W., Gaudzinski-Windheuser S., Bruhn K. C., Tost H., Sirocko F., Crutzen P. J. 2013. The Palaeoanthropocene: the beginnings of anthropogenic environmental change. Anthropocene 3: 83–88.

Fong M.W., Gao H., Wong J.Y., et al.2020. Nonpharmaceutical measures for pandemic influenza in nonhealthcare settings— social distancing measures. Emerg Infect Dis 2020; Published online Feb 6. DOI:10.3201/eid2605.190995.

Fraser C., Riley S., Anderson R.M., Ferguson N.M. 2004. Factors that make an infectious disease outbreak controllable. Proc Natl Acad Sci USA; 101: 6146–51.

Friedman M.S., Powell K.E., Hutwagner L., Graham L.M., Teague W.G.2001. Impact of changes in transportation and commuting behaviors during the 1996 Summer Olympic Games in Atlanta on air quality and childhood asthma. JAMA; 285: 897–905.

Fröohlich-Nowoisky J., Kampf C.J., Weber B., Huffman J.A., Pöschl U. 2016. Bioaerosols in the Earth system: climate, health, and ecosystem interactions. Atmos. Res. 182, 346–376.

Funk S., Ciglenecki I., Tiffany A., et al. 2017. The impact of control strategies and behavioural changes on the elimination of Ebola from Lofa County, Liberia. Philos Trans R Soc Lond B Biol Sci 2017; 372: 20160302.

Gao, J.F., Fan, X.Y., Li, H.Y., Pan, K.L., 2017. Airborne bacterial communities of PM2.5 in Beijing-Tianjin-Hebei megalopolis, China as revealed by Illumina MiSeq sequencing: a case study. Aerosol. Air. Qual. Res. 17.

Ghio A.J., M.S. Carraway, M.C. Madden, 2012. Composition of air pollution particles and oxidative stress in cells, tissues, and living systems, J. Toxicol. Environ. Health B Crit. Rev. 15 (1) (2012) 1–21.

Glasser J.W., Hupert N., McCauley M.M., Hatchett R. 2011. Modeling and public health emergency responses: lessons from SARS. Epidemics 2011; 3: 32–37.

Glencross Drew A., Tzer-Ren Ho, Nuria Camina, Hawrylowicz Catherine M., Pfeffer P. E. 2020. Air pollution and its effects on the immune system, Free Radical Biology and Medicine, in press. https://doi.org/10.1016/j.freeradbiomed.2020.01.179

Glikson A. 2013. Fire and human evolution: The deep-time blueprints of the Anthropocene. Anthropocene 3, 89–92.

Gorse G.J., O’Connor T.Z., Young S.L., Habib M.P., Wittes J., Neuzil K.M., et al. 2006. Impact of a winter respiratory virus season on patients with COPD and association with influenza vaccination. Chest 2006; 130:1109–16.

Grant W. B. 2002. An ecologic study of dietary and solar Ultraviolet-B links to breast carcinoma mortality rates. Cancer 94(1):272–281

Groulx N., Urch B., Duchaine C., Mubareka S., Scott J. A. 2018. The Pollution Particulate Concentrator (PoPCon): A platform to investigate the effects of particulate air pollutants on viral infectivity, Science of the Total Environment 628–629 (2018) 1101–1107, https://doi.org/10.1016/j.scitotenv.2018.02.118

Guo L., aJiaLuoaManYuanaYapingHuangaHuanfengShenbTongwenLib 2019. The influence of urban planning factors on PM2.5 pollution exposure and implications: A case study in China based on remote sensing, LBS, and GIS data. Science of The Total Environment Volume 659, 1 April 2019, Pages 1585–1596, https://doi.org/10.1016/j.scitotenv.2018.12.448

Hao J., Zhiyi Yang, Shuqiong Huang, Wenwen Yang, Zhongmin Zhud, Liqiao Tian, Yuanan Luf, Hao Xiang, Suyang Liu 2019. The association between short-term exposure to ambient air pollution and the incidence of mumps in Wuhan, China: A time-series study. Environmental Research 177, 108660, https://doi.org/10.1016/j.envres.2019.108660

Hellewell J., Abbott S., Gimma A., Bosse N. I., Jarvis C. I., Russell T. W., Munday J.D., Kucharski A.J., Edmunds W. J., Centre for the Mathematical Modelling of Infectious Diseases COVID-19 Working Group, Sebastian Funk, Eggo R. M 2020. Feasibility of controlling COVID-19 outbreaks by isolation of cases and contacts, Lancet Glob Health 2020, https://doi.org/10.1016/S2214-109X(20)30074-7

Hoang T.T.T., Nguyen V.N., Dinh N.S., et al. 2019. Active contact tracing beyond the household in multidrug resistant tuberculosis in Vietnam: a cohort study. BMC Public Health 2019; 19: 241.

Hoek G., Krishnan R.M., Beelen R., Peters A., Ostro B., Brunekreef B., Kaufman J.D., 2013 Dec. Long-term air pollution exposure and cardiorespiratory mortality: a review. Environ. Health 12 (1), 43.

Il meteo 2020. Medie e totali mensili. https://www.ilmeteo.it/portale/medie-climatiche (Accessed March 2020).

Istituto Superiore Sanità, 2020. Nuovo coronavirus SARS-CoV-2. Caratteristiche dei pazienti deceduti positivi a COVID-19 in Italia, https://www.epicentro.iss.it/coronavirus/sars-cov-2-decessi-italia (Accessed 1 April, 2020).

Jalaludin B.B., O’Toole B.I., Leeder S.R., 2004. Acute effects of urban ambient air pollution on respiratory symptoms, asthma medication use, and doctor visits for asthma in a cohort of Australian children. Environ. Res. 95, 32–42. http://dx.doi.org/10.1016/S0013-9351(03)00038-0.

Jansen A.G.S.C., Sanders E.A.M., Van Der Ende A., Van Loon A.M., Hoes A.W., Hak E. 2008. Invasive pneumococcal and meningococcal disease: association with influenza virus and respiratory syncytial virus activity? Epidemiol Infect; 136:1448e54.

Jaspers I., Ciencewicki J.M., Zhang W.L., Brighton L.E., Carson J.L., Beck M.A., et al. 2005. Diesel exhaust enhances influenza virus infections in respiratory epithelial cells. Toxicol Sci; 85:990–1002.

Jin, L., Luo, X., Fu, P., Li, X., 2017. Airborne particulate matter pollution in urban China: a chemical mixture perspective from sources to impacts. Natl. Sci. Rev. 593, 610.

Johns Hopkins Center for System Science and Engineering, 2020. Coronavirus COVID-19 Global Cases, https://gisanddata.maps.arcgis.com/apps/opsdashboard/index.html#/bda7594740fd40299423467b48e9ecf6 (Accessed in April 2020).

Jones A.M., Harrison, R.M., 2004. The effects of meteorological factors on atmospheric bio aerosol concentrations-a review. Sci. Total Environ. 326, 151e180

Jung C.R., Hsieh, H.Y., Hwang, B.F., 2017. Air pollution as a potential determinant of rheumatoid arthritis: a population-based cohort study in Taiwan. Epidemiology 28, S54–S59.

Kampa M., Castanas E., 2008. Human health effects of air pollution. Environ. Pollut. 151 (2), 362–367.

Kang M., Song T., Zhong H., et al. 2016. Contact tracing for imported case of Middle East respiratory syndrome, China, 2015.Emerging Infect Dis 2016; 22: 9.

Kim P.E., Musher D.M., Glezen W.P., Rodriguez-Barradas M.C., Nahm W.K., Wright C.E. 1996. Association of invasive pneumococcal disease with season, atmospheric conditions, air pollution, and the isolation of respiratory viruses. Clin Infect Dis 1996; 22:100e6.

Ko F.W.S., Chan P.K.S., Chan M.C.H., To K.W., Ng S.S.S., Chau S.S.L., et al. 2007. Viral etiology of acute exacerbations of COPD in Hong Kong. Chest; 132:900–8.

Ko F.W.S., Tam W., Wong T.W., Chan D.P.S., Tung A.H., Lai C.K.W. et al. 2007a. Temporal relationship between air pollutants and hospital admissions for chronic obstructive pulmonary disease in Hong Kong. Thorax 2007; 62:779e84.

Kucharski A.J., Camacho A., Checchi F. et al. 2015. Evaluation of the benefits and risks of introducing Ebola community care centers, Sierra Leone. Emerg Infect Dis 2015; 21: 393–99.

Kucharski Adam J, Timothy W Russell, Charlie Diamond, Yang Liu, John Edmunds, Sebastian Funk, Rosalind M Eggo, on behalf of the Centre for Mathematical Modelling of Infectious Diseases COVID-19 working group* 2020. Early dynamics of transmission and control of COVID-19: a mathematical modelling study Lancet Infect Dis 2020 Published Online March 11, 2020 https://doi.org/10.1016/S1473-3099(20)30144-4. See Online/Commenthttps://doi.org/10.1016/S1473-3099(20)30161-4

Langrish J. P., Mills N. L. 2014. Air pollution and mortality in Europe, Lancet, vol. 383, http://dx.doi.org/10.1016/S0140-6736(13)62570-2

Legambiente(2019)Mal’aria2019,ilrapportoannualesull’inquinamentoatmosfericonellecittàitaliane. https://www.legambiente.it/malaria-2019-il-rapporto-annuale-annuale-sullinquinamento-atmosferico-nelle-citta-italiane/ (Accessed March 2020)

Lewtas J. 2007. Air pollution combustion emissions: Characterization of causative agents and mechanisms associated with cancer, reproductive, and cardiovascular effects. Mutation Research, vol. 636, Issues 1–3, pp. 95–133, https://doi.org/10.1016/j.mrrev.2007.08.003.

Li J., Sun S., Tang R., Qiu H., Huang Q., Mason T.G., Tian L. 2016. Major air pollutants and risk of COPD exacerbations: a systematic review and meta-analysis, Int. J. Chronic Obstr. Pulm. Dis. 11, 3079–3091.

Li Q., Guan X., Wu P., et al. 2020. Early transmission dynamics in Wuhan, China, of novel coronavirus-infected pneumonia. N Engl J Med 2020; Published online Jan 29. DOI:10.1056/NEJMoa2001316.

Liao C.-M., Nan-Hung Hsieh, Chia-Pin Chio 2011. Fluctuation analysis-based risk assessment for respiratory virus activity and air pollution associated asthma incidence. Science of the Total Environment 409, 3325–3333, doi: 10.1016/j.scitotenv.2011.04.056

Lim H.S. et al. 2006. Cancer survival is dependent on season of diagnosis and sunlight exposure. Int J Can 119:1530–1536

Linstone H. A. 1999. Decision making for technology executives, Artech House, Boston-London

Liu H., Zhang X., Zhang H., Yao X., Zhou M., Wang J., et al., 2018. Effect of air pollution on the total bacteria and pathogenic bacteria in different sizes of particulate matter. Environ. Pollut. 233, 483–493.

Liu M., Huang Y., Ma Z., Jin Z., Liu X., Wang H., et al., 2017. Spatial and temporal trends in the mortality burden of air pollution in China: 2004-2012. Environ. Int. 98, 75–81.

Ma Y., Zhao Y., Liu J., He X., Wang B., Fu S., Yan J., Niu J., Zhou J., Luo B. 2020. Effects of temperature variation and humidity on the death of COVID-19 in Wuhan, China, Science of The Total Environment,138226, https://doi.org/10.1016/j.scitotenv.2020.138226.

Manuell M-E, Cukor J. 2011. Mother Nature versus human nature: public compliance with evacuation and quarantine. Disasters 2011; 35: 417–42.

McCullers JA.2006. Insights into the interaction between influenza virus and pneumococcus. Clin Microbiol Rev; 19:571e82.

Medina-Ramón M., Zanobetti A., Schwartz J. 2006. The effect of ozone and PM10 on hospital admissions for pneumonia and chronic obstructive pulmonary disease: a national multicity study. Am J Epidemiol; 163:579e88.

MinisterodellaSalute2020.Covid-19-SituazioneinItalia. http://www.salute.gov.it/portale/nuovocoronavirus/dettaglioContenutiNuovoCoronavirus.jsp?lingua=italiano&id=5351&area=nuovoCoronavirus&menu=vuoto (Accessed March 2020)

Mochitate K., Katagiri K., Miura T. 2001. Impairment of microbial billing and superoxide-producing activities of alveolar macrophages by a low level of ozone. J Health Sci 2001; 47: 302–09.

Murdoch D. R., Jennings Lance C. 2009. Association of respiratory virus activity and environmental factors with the incidence of invasive pneumococcal disease, Journal of Infection, 58, 37–46, doi: 10.1016/j.jinf.2008.10.011

Murphy K.R., Eivindson A., Pauksens K., Stein W.J., Tellier G., Watts R. et al. 2000. Efficacy and safety of inhaled zanamivir for the treatment of influenza in patients with asthma or chronic obstructive pulmonary disease — a double-blind, randomised, placebo controlled, multicentre study. Clin Drug Invest; 20:337–49.

Myllykangas-Luosujäarvi R., Aho K., Kautiainen H., Isomäki H., 1995. Shortening of life span and causes of excess mortality in a population-based series of subjects with rheumatoid arthritis. Clin. Exp. Rheumatol. 13 (2), 149–153.

nCoV-2019 Data Working Group. 2020. Epidemiological data from the nCoV-2019 outbreak: early descriptions from publicly Available data. 2020. http://virological.org/t/epidemiological-data-from-the-ncov-2019-outbreak-early-descriptions-from-publicly-Available-data/337(Accessed Feb 13, 2020).

Nel A. 2005. Air pollution-related illness: effects of particles. Science 2005; 308:804–6.

Nenna R., Evangelisti M., Frassanito A., Scagnolari C., Pierangeli A., Antonelli G., Nicolai A., Arima S., Moretti C., Papoff P., Villa M. P., Midulla F. 2017. Respiratory syncytial virus bronchiolitis, weather conditions and air pollution in an Italian urban area: An observational study. Environmental Research 158 (2017) 188–193, http://dx.doi.org/10.1016/j.envres.2017.06.014

Neu U., Mainou B.A. 2020. Virus interactions with bacteria: Partners in the infectious dance. PLoS Pathog 16(2): e1008234. https://doi.org/10.1371/journal.ppat.1008234

Oh E.-Y., Ansell C., Nawaz H., Yang C.-H., Wood P. A., Hrushesky W.J. M. 2010. Global breast cancer seasonality, Breast Cancer Res Treat (2010) 123:233–243. DOI 10.1007/s10549-009-0676-7

Orellano P., Quaranta N., Reynoso J., Balbi B., Vasquez J. 2017. Effect of outdoor air pollution on asthma exacerbations in children and adults: systematic review and multilevel meta-analysis, PloS One 12 (3) (2017) e0174050.

Peak C.M., Childs L.M., Grad Y.H., Buckee C.O. 2017. Comparing nonpharmaceutical interventions for containing emerging epidemics. Proc Natl Acad Sci USA 2017; 114: 4023–28.

Pike A. 2009. De-Industrialization. Elsevier Ltd. All rights reserved.

Pope C.A. 1989. Respiratory disease associated with community air pollution and a steel mill. Utah Val Am J Public Health; 79: 623–28.

Pope C.A. 1996. Particulate pollution and health: a review of the Utah valley experience. J Expo Anal Environ Epidemiol; 6: 23–34.

Pope C.A. Ezzati M., Dockery D.W. 2009. Fine-particulate air pollution and life expectancy in the United States. N Engl J Med 2009; 360: 376–86.

Prem K., Liu Y., Russell T. W., Kucharski A.J., Eggo R. M., Davies N. et al., 2020. The effect of control strategies to reduce social mixing on outcomes of the COVID-19 epidemic in Wuhan, China: a modelling study, The Lancet Public Health, March 25, 2020 https://doi.org/10.1016/S2468-2667(20)30073-6

Public Health England, 2019. MERS-CoV close contact algorithm. Public health investigation and management of close contactsofMiddleEastRespiratoryCoronavirus(MERS-CoV)cases(v1729January2019). 2019.https://assets.publishing.service.gov.uk/government/uploads/system/uploads/attachment_data/file/776218/MERS-CoV_Close_contacts_algorithm.pdf (Accessed Feb 6, 2020).

PublicHealthEngland,2020.Novelcoronavirus(2019-nCoV)–whatyouneedtoknow.2020. https://publichealthmatters.blog.gov.uk/2020/01/23/wuhan-novel-coronavirus-what-you-need-to-know/ (Accessed Jan 31, 2020).

Quilty B., Clifford S. CCMID nCoV working group, Flasche S, Eggo RM. 2020. Effectiveness of airport screening at detecting travelersinfectedwith2019-nCoV.2020.

https://cmmid.github.io/ncov/airport_screening_report/airport_screening_preprint_2020_01_30.pdf (Accessed Feb 5, 2020).

Raaschou-Nielsen O., Andersen Z.J., Beelen R., et al. Air pollution and lung cancer incidence in 17 European cohorts: prospective analyses from the European Study of Cohorts for Air Pollution Effects (ESCAPE). Lancet Oncol 2013; 14: 813–22.

Rahman I., MacNee W. 2000. Oxidative stress and regulation of glutathione in lung inflammation. Eur Respir J; 16: 534–54.

Riley S, Fraser C, Donnelly CA, et al. 2003. Transmission dynamics of the etiological agent of SARS in Hong Kong: impact of public health interventions. Science 2003; 300: 1961–66.

Riou J., Althaus C.L. 2020. Pattern of early human-to-human transmission of Wuhan 2019 novel coronavirus (2019-nCoV), December 2019 to January 2020. Euro Surveill 2020; 25: 2000058.

Ségala C., Poizeau D., Mesbah M., Willems S., Maidenberg M., 2008. Winter air pollution and infant bronchiolitis in Paris. Environ. Res. 106, 96–100. http://dx.doi.org/10.1016/j.envres.2007.05.003.

Shankardass K., McConnell R., Jerrett M., Milam J., Richardson J., Berhane K. 2009. Parental stress increases the effect of traffic-related air pollution on childhood asthma incidence. Proc Natl Acad Sci USA; 106:12406–1

Shepherd A., Mullins J. T. 2019. Arthritis diagnosis and early-life exposure to air pollution, Environmental Pollution 253 (2019) 1030–1037, https://doi.org/10.1016/j.envpol.2019.07.054

Simoni M., Baldacci S., Maio S., Cerrai S., Sarno G., Viegi G., 2015. Adverse effects of outdoor pollution in the elderly. J. Thorac. Dis. 7, 34–45. http://dx.doi.org/10.3978/j.issn.2072-1439.2014.12.10.

Smets W., Morett, S., Denys S., Lebeer S. 2016. Airborne bacteria in the atmosphere: presence, purpose, and potential. Atmos. Environ. 139, 214e221

Sun, Y., Xu, S., Zheng, D., Li, J., Tian, H., Wang, Y., 2018. Effects of haze pollution on microbial community changes and correlation with chemical components in atmospheric particulate matter. Sci. Total Environ. 637e638, 507.

Swanson K.C., Altare C., Wesseh C.S., et al. 2018. Contact tracing performance during the Ebola epidemic in Liberia, 2014– 2015. PLoS Negl Trop Dis 2018; 12: e0006762.

Talbot T.R., Poehling K.A., Hartert T.V., Arbogast P.G., Halasa N.B., Edwards K.M., et al.2005. Seasonality of invasive pneumococcal disease: temporal relation to documented influenza and respiratory syncytial viral circulation. Am J Med;118: 285–91.

The Italian National Institute of Statistics (ISTAT, 2020). Popolazione residente al 1° gennaio, http://dati.istat.it/Index.aspx?DataSetCode=DCIS_POPRES1

U. S. Census Bureau 2020. U.S. and World Population Clock, https://www.census.gov/popclock/ (Accessed April, 2020)

Van Leuken, J.P.G., Swart A.N., Havelaar A.H., Van Pul A., Van der Hoek W., Heederik D. 2016. Atmospheric dispersion modelling of bio aerosols that are pathogenic to humans and livestock - a review to inform risk assessment studies. Microb. Risk. Anal. 1, 19–39.

Vandini S., Bottau P., Faldella G., Lanari L. 2015. Immunological, viral, environmental, and individual factors modulating lung immune response to respiratory syncytial virus. Biomed. Res. Int. 2015, 875723. http://dx.doi.org/10.1155/2015/875723.

Vandini S., Corvaglia L., Alessandroni R., Aquilano G., Marsico C., Spinelli M., Lanari M., Faldella G., 2013. Respiratory syncytial virus infection in infants and correlation with meteorological factors and air pollutants. Ital. J. Pediatr. 39, 1. http://dx.doi.org/10.1186/1824-7288-39-1.

Wang C., Horby P.W., Hayden F.G., Gao G.F. 2020. A novel coronavirus outbreak of global health concern. Lancet 2020; 395: 470–73

Wang G., Zhang R., Gomez M.E., Yang L., Zamora M.L., Hu M., Lin Y., Peng J., Guo S., Meng J., Li J. 2016. Persistent sulfate formation from London Fog to Chinese haze. Proc. Natl. Acad. Sci. 113 (48), 13630e13635.

Ward DJ, Ayres JG. 2004. Particulate air pollution and panel studies in children: a systematic review. Occup Environ Med; 61: e13.

Wei M., Houfeng Liu, Jianmin Chen, Caihong Xu, Jie Li, Pengju Xu, Ziwen Sun 2020. Effects of aerosol pollution on PM2.5-associated bacteria in typical inland and coastal cities of northern China during the winter heating season, Environmental Pollution 262 (2020) 114188, https://doi.org/10.1016/j.envpol.2020.114188

Wei, K., Zou, Z., Zheng, Y., Li, J., Shen, F., Wu, C.Y., et al., 2016. Ambient bio aerosol particle dynamics observed during haze and sunny days in Beijing. Sci. Total Environ. 550, 751e759.

Weinmayr G., Romeo E., De Sario M., Weiland S.K., Forastiere F. 2010. Short term effects of PM10 and NO2 on respiratory health among children with asthma or asthma-like symptoms: a systematic review and meta-analysis. Environ Health Perspect; 118:449–57.

Wells C. R., Sah P., Moghadas S. M., Pandey A., Shoukat A., Wang Y., Wang Z., Meyers L. A., Singer B. H., Galvani A. P.2020. Impact of international travel and border control measures on the global spread of the novel 2019 coronavirus outbreak. Proceedings of the National Academy of Sciences Mar 2020, 202002616; DOI: 10.1073/pnas.2002616117

WHO 2019. Coronavirus disease 2019 (COVID-19). Situation report 24. February 13, 2020. Geneva: World Health Organization, 2020.

WHO2020.Novelcoronavirus(2019-nCoV)situationreport16.n-reports/20200205-sitrep-16- ncov.pdf?sfvrsn=23af287f_2 (Accessed Feb 5, 2020).

WHO2020a.Novelcoronavirus(2019-nCoV)situationreport2.WorldHealthOrganization,2020. https://www.who.int/docs/defaultsource/coronaviruse/situation-reports/20200122-sitrep-2-2019-ncov.pdf?sfvrsn=4d5bcbca_2 (Accessed Jan 22, 2020).

WHO 2020b. Implementation and management of contact tracing for Ebola virus disease. World Health Organization. 2015.http://www.who.int/csr/resources/publications/ebola/contacttracing/en/ (Accessed Feb 4, 2020).

WHO 2020c. Who Director-General’s opening remarks at the media briefing on COVID-19. March 3, 2020. https://www.who.int/dg/speeches/detail/whodirector-general-s-opening-remarks-at-the-media-briefing-on-covid-19---3-march-2020 (Accessed March 6, 2020).

Wilder-Smith A., Chiew C.J., Lee V.J. 2020. Can we contain the COVID-19 outbreak with the same measures as for SARS? Lancet Infect Dis 2020; Published online March 5. https://doi.org/10.1016/S1473-3099(20)30129-8.

Wong C.M., Yang L., Thach T.Q., Chau P.Y.K., Chan K.P., Thomas G.N., et al. 2009. Modification by influenza on health effects of air pollution in Hong Kong. Environ Health Perspect; 117:248–53.

Wooding D.J., Ryu M.H., Huls A., Lee A.D., Lin D.T.S., Rider C.F., Yuen A.C.Y., Carlsten C. 2019. Particle depletion does not remediate acute effects of traffic-related air pollution and allergen. A randomized, double-blind crossover study, Am. J. Respir. Crit. Care Med. 200 (5) (2019) 565–574.

Wu J.T., Leung K., Leung G.M. 2020. Now casting and forecasting the potential domestic and international spread of the 2019-nCoV outbreak originating in Wuhan, China: a modelling study. Lancet 2020; 395: 689–97.

Xie Z.S., Fan C.L., Lu R., Liu P.X., Wang B.B., Du S.L., et al. 2018. Characteristics of ambient bio aerosols during haze episodes in China: a review. Environ. Pollut. 243, 1930e1942.

Xu B., Kraemer Moritz U. G. 2020. Open access epidemiological data from the COVID-19 outbreak, The Lancet Infectious Diseases, https://doi.org/10.1016/S0140-6736(20)30371, Available online 19 February 2020

Yang Z., Jiayuan Hao, Shuqiong Huang, Wenwen Yang, Zhongmin Zhu, Liqiao Tian, Yuanan Lu, Hao Xiang, Suyang Liu 2020. Acute effects of air pollution on the incidence of hand, foot, and mouth disease in Wuhan, China. Atmospheric Environment 225 (2020) 117358, https://doi.org/10.1016/j.atmosenv.2020.117358

Yao X., Ye F., Zhang M., et al. In vitro antiviral activity and projection of optimized dosing design of hydroxychloroquine for the treatment of severe acute respiratory syndrome coronavirus 2 (SARS-CoV-2). Clin Infect Dis 2020; Published online March 9. DOI:10.1093/cid/ciaa237.

Zhai Y., Li X., Wang T., Wang B., Li C., Zeng G., 2018. A review on airborne microorganisms in particulate matters: composition, characteristics and influence factors. Environ. Int. 113, 74e90.

Zhang Q., Zheng Y., Tong D., Shao M., Wang S., Zhang Y., Xu X., Wang J., He H., Liu W., Ding Y., Lei Y., Li J., Wang Z., Zhang X., Wang Y., Cheng J., Liu Y., Shi Q., Yan L., Geng G., Hong C., Li M., Liu F., Zheng B., Cao J., Ding A., Gao J., Fu Q., Huo J., Liu B., Liu Z., Yang F., He K., Hao J. 2019a. Drivers of improved PM2.5 air quality in China from 2013 to 2017, Proceedings of the National Academy of Sciences Dec, 116 (49) 24463–24469; DOI: 10.1073/pnas.1907956116

Zhang T., Li X., Wang M., Chen H., Yao M., 2019. Microbial aerosol chemistry characteristics in highly polluted air. Sci. China Chem. 62 https://doi.org/10.1007/s11426-11019-19488-11423.

Zhang Y., Ding A., Mao H., We, N., Zhou D., Liu L. et al., 2016. Impact of synoptic weather patterns and inter-decadal climate variability on air quality in the North China Plain during 1980e2013. Atmos. Environ. 124, 119e128

Zheng X.Y., H. Ding, L.N. Jiang, S.W. Chen, J.P. Zheng, M. Qiu, Y.X. Zhou, Q. Chen, W.J. Guan, 2015. Association between air pollutants and asthma emergency room visits and hospital admissions in time series studies: a systematic review and meta-analysis, PloS One 10 (9) (2015) e0138146

Zhong J., Zhang X., Dong Y., Wang Y., Wang J., Zhang Y., et al., 2018. Feedback effects of boundary-layer meteorological factors on explosive growth of PM2.5 during winter heavy pollution episodes in Beijing from 2013 to 2016. Atmos. Chem. Phys. 18, 247e258.

Zhou W. et al. 2005 Vitamin D is associated with improved survival in early-stage non-small cell lung cancer patients. Cancer Epidemiol Biomarkers Prev 14(10):2303–2309

Zhu N., Zhang D., Wang W., et al. 2020. A novel coronavirus from patients with pneumonia in China, 2019. N Engl J Med 2020; Published Feb 20. DOI:10.1056/NEJMoa2001017.

Zhu Y., Xie J. 2020. Association between ambient temperature and COVID-19 infection in 122 cities from China, Science of the Total Environment, https://doi.org/10.1016/j.scitotenv.2020.138201

